# Clinical syndromes linked to biallelic germline variants in *MCM8* and *MCM9*

**DOI:** 10.1101/2024.10.30.24315828

**Authors:** Noah C. Helderman, Ting Yang, Claire Palles, Diantha Terlouw, Hailiang Mei, Ruben H.P. Vorderman, Davy Cats, Marcos Díaz Gay, Marjolijn C.J. Jongmans, Ashwin Ramdien, MCM8-MCM9 study group, Mariano Golubicki, Marina Antelo, Laia Bonjoch, Mariona Terradas, Laura Valle, Ludmil B. Alexandrov, Hans Morreau, Tom van Wezel, Sergi Castellví-Bel, Yael Goldberg, Maartje Nielsen, Irma van de Beek, Thomas F. Eleveld, Andrew Green, Frederik J. Hes, Marry M. van den Heuvel-Eibrink, Annelore Van Der Kelen, Sabine Kliesch, Roland P. Kuiper, Inge M.M. Lakeman, Lisa E.E.L.O. Lashley, Leendert H.J. Looijenga, Manon S. Oud, Johanna Steingröver, Yardena Tenenbaum-Rakover, Carli M. Tops, Frank Tüttelmann, Richarda M. de Voer, Dineke Westra, Margot J. Wyrwoll

## Abstract

**Background:** *MCM8* and *MCM9* are newly proposed cancer predisposition genes, linked to polyposis and early-onset cancer, in addition to their association with hypogonadism. Given the uncertain range of phenotypic manifestations and unclear cancer risk estimates, this study aimed to delineate the molecular and clinical characteristics of individuals with biallelic germline *MCM8*/*MCM9* variants.

**Methods:** Population allele frequencies and biallelic variant carrier frequencies were calculated using data from gnomAD, and a variant enrichment analysis was conducted across multiple cancer and non-cancer phenotypes using data from the 100K Genomes Project and the 200K exome release of the UK Biobank. A case series was conducted, including previously reported variant carriers with and without updated clinical data and newly identified carriers through the European Reference Network (ERN) initiative for rare genetic tumor risk syndromes (GENTURIS). Additionally, mutational signature analysis was performed on tumor data from our case series and publicly available datasets from the Hartwig Medical Foundation and TCGA Pan-Cancer Atlas to identify mutational signatures potentially associated with MCM8/MCM9 deficiency.

**Results:** Predicted loss of function and missense variants in *MCM8* (1.4 per 100,000 individuals) and *MCM9* (2.5 per 100,000 individuals) were found to be rare in gnomAD. However, biallelic *MCM9* variants showed significant enrichment in cases from the 100K Genomes Project compared to controls for colonic polyps (odds ratio (OR) 6.51, 95% confidence interval (CI) 1.24–34.11; *P* = 0.03), rectal polyps (OR 8.40, 95% CI 1.28–55.35; *P* = 0.03), and gastric cancer (OR 27.03, 95% CI 2.93– 248.5; *P* = 0.004). No significant enrichment was found for biallelic *MCM8* variant carriers or in the 200K UK Biobank. In our case series, which included 26 biallelic *MCM8* and 28 biallelic *MCM9* variant carriers, we documented polyposis, gastric cancer, and early-onset colorectal cancer in 6, 1, and 6 biallelic *MCM9* variant carriers, respectively, while these phenotypes were not observed in biallelic *MCM8* variant carriers. Additionally, our case series indicates that, beyond hypogonadism—which was present in 23 and 26 of the carriers, respectively—biallelic *MCM8* and *MCM9* variants are associated with early-onset germ cell tumors (occurring before age 15) in 2 *MCM8* and 1 *MCM9* variant carriers. Tumors from *MCM8*/*MCM9* variant carriers with available germline sequencing data predominantly displayed clock-like mutational processes (single base substitution signatures 1 and 5), with no evidence of signatures associated with DNA repair deficiencies.

**Discussion:** Our data indicates that biallelic *MCM9* variants are associated with polyposis, gastric cancer, and early-onset CRC, while both biallelic *MCM8* and *MCM9* variants are linked to hypogonadism and the early development of germ cell tumors. These findings underscore the importance of including *MCM8*/*MCM9* in diagnostic gene panels for certain clinical contexts and suggest that biallelic carriers may benefit from cancer surveillance.

## Introduction

The identification of cancer predisposition syndromes plays a crucial role in preventing and surveilling malignancies at an early stage in affected individuals. Nevertheless, a significant proportion of familial cancer cases lack a clear explanation.^1^ This poses challenges in developing personalized surveillance programs and highlights the urgency of exploring and identifying novel cancer predisposition genes.

The minichromosome maintenance 8 homologous recombination repair factor (*MCM8*; NM_032485.6, ENST00000610722.4, OMIM 608187) and minichromosome maintenance 9 homologous recombination repair factor (*MCM9*; NM_017696.3, ENST00000619706.5, OMIM 610098) genes are two recently suggested cancer predisposition genes.^2–4^ The proteins encoded by these genes form a helicase hexameric complex that is likely involved in DNA replication and the initiation of DNA replication^5–9^, meiosis^7, 10–13^, homologous recombination^14–20^ and mismatch repair (MMR).^4, 19, 21^

Following their significant association with primary ovarian insufficiency (POI; HP:0008209) ^2–4, 22–40^, biallelic germline variants of *MCM8/MCM9* were first linked to cancer in several families with polyposis (HP:0200063) and early-onset colorectal cancer (CRC; HP:0003003).^2–4^ Subsequently, there have been reports of individuals with CRC carrying a monoallelic *MCM8/MCM9* variant^2–4^, as well as reports describing mono-and biallelic germline *MCM8/MCM9* variants in patients with other nonmalignant pathologies, including short stature (HP:0004322)^29, 34, 35, 38, 39^, delayed puberty (HP:0000823)^22, 23, 26, 28, 33, 38–40^, hypothyroidism (HP:0000821)^22, 28^, and absent or infantile uteri/ovaries.^22, 23, 26, 27, 29, 31, 33, 35, 37–40^

Due to the limited number of families with biallelic germline *MCM8/MCM9* variants described so far, the complete spectrum of phenotypic manifestations and accurate cancer risk estimates remain uncertain. As a result, the incorporation of the *MCM8/MCM9* genes into diagnostic gene panels is not widespread, and the respective syndrome(s) associated with both genes could easily be missed. This study, therefore, sought to delineate the molecular and clinical features of biallelic germline *MCM8/MCM9* variants and to establish recommendations for the clinical management of variant carriers.

## Methods

### Ethical statement

This study was approved by the local IRB and biobank committee of the Leiden University Medical Center in The Netherlands (protocol B18.007). Storage and management of clinical and molecular data and patient samples from our case series was supervised by the Leiden University Medical Center in The Netherlands. Patient samples were handled according to the medical ethical guidelines described in the Code of Conduct for responsible use of human tissue in the context of health research (Federation of Dutch Medical Scientific Societies). Samples were coded/anonymized, and all patients provided written informed consent for the use of tissue and data. The patient and sample IDs used in this study cannot be used to reveal participant identities by anyone outside the research team.

### Population-based cohorts

#### Estimation of population allele and biallelic carrier frequencies in gnomAD v.2.1.1

The gnomAD (v.2.1.1, containing 141,456 samples; https://gnomad.broadinstitute.org/) database was accessed in May 2023 to estimate the allele frequencies of *MCM8/MCM9* variants across multiple populations. Predicted loss of function variants (pLoF; including splice acceptor, splice donor, frameshift, and stop gained variants) and missense variants, as classified based on the Ensembl Variant Effect Predictor (VEP) consequence^41^, were analyzed separately for both genes (**Figure 1**). Allele frequencies are presented as # of cases per 100,000 persons, unless otherwise specified.

**Figure 1.**
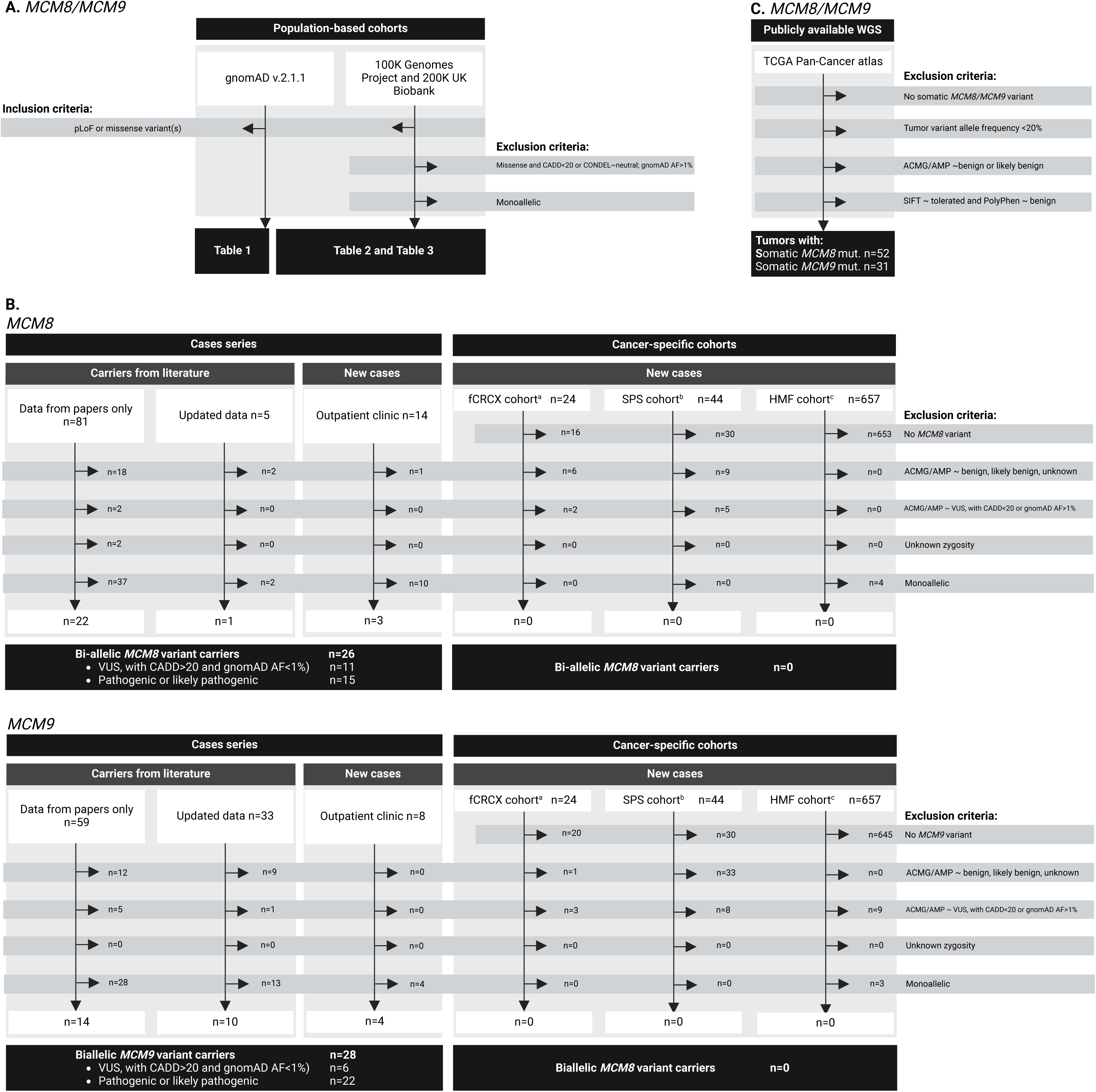
Flow chart of study approach. Pathogenicity-based filtering of (**A**) population-based cohorts, (**B**) our case series and cancer-specific cohorts, and (**C**) the TCGA Pan-Cancer atlas dataset. ^a^fCRCX cohort comprised 24 CRC-affected members of 16 Amsterdam-positive non-polyposis CRC families; ^b^SPS cohort comprised 44 unrelated serrated polyposis families; ^c^HMF cohort comprised 632 metastasized CRCs and 25 metastasized ECs. Tumors from the TCGA Pan-Cancer Atlas were selected based on the presence of somatic *MCM8*/*MCM9* variants and are not related to germline variant carriers. *ACMG/AMP, American College of Medical Genetics and Genomics; AF, allele frequency; CADD, Combined Annotation-Dependent Depletion; CRC, colorectal cancer; EC, endometrial cancer; pLoF, predicted loss of function; VUS, variant of uncertain significance*.

The gnomAD (v.2.1.1, exomes only, containing 125,748 samples; https://gnomad.broadinstitute.org/) database was assessed in May 2023 to estimate the biallelic carrier frequencies of *MCM8/MCM9* variants. These carriers could be either homozygous or compound heterozygous for the *MCM8/MCM9* variant(s). For compound heterozygous variant carriers, we included individuals who carried two heterozygous variants in *trans* (on different copies of the genes), while individuals with two heterozygous variants in *cis* (on the same copy of the gene) were excluded from the analysis. Phasing of the variants was predicted by the gnomAD variant co-occurrence tool (gnomad.broadinstitute.org/variant-cooccurrence).

#### Identification of carriers and variant enrichment analysis in 200K UK Biobank and 100K Genomes Project datasets

Germline variants in *MCM8* and *MCM9* were identified from the 100,000 Genomes Project (project code 1142, version 17) and the 200K exomes release of the UK Biobank (project code 86977, released on November 17, 2021). Variants were annotated using Variant Effect Predictor (VEP) v.107.^41^ We retained missense variants with a Combined Annotation-Dependent Depletion (CADD)^42^ score ≥20 and a deleterious Condel score^43^, as well as pLoF variants (including splice acceptor, splice donor, frameshift, and stop gained variants), provided their allele frequency was <1% in gnomAD v2.1.1 (**Figure 1**). The impact of variants on the canonical transcripts was reported for *MCM8* (ENST00000610722.4) and *MCM9* (ENST00000619706.5).

The International Classification of Diseases 10th Revision (ICD10) codes from participants’ diagnosis information (Participant Explorer in 100K Genomes Project, field ID 41270 in 200K UK Biobank), along with the International Classification of Diseases for Oncology (ICD-O) codes obtained from cancer histology and behavior fields (field ID 40011 and 40012 in 200K UK Biobank), as shown in **Supplementary Table 1**, were searched to identify participants with phenotypes associated with *MCM8/MCM9* variants, as selected based on literature^44^ as well as our case series.

We conducted case-control tests to assess whether potentially pathogenic biallelic (homozygous or compound heterozygous) *MCM8*/*MCM9* variants were enriched in participants with the phenotypes of interest compared to a control cohort. A total of 15,091 controls were identified from the 100K Genomes Project dataset and 90,897 from the 200K UK Biobank dataset. Controls were selected based on the absence of personal or family history of common cancers and any phenotypes listed in **Supplementary Table 1**. To account for differences in age and ethnicity between cases and controls, association testing was performed using PLINK v1.9, adjusting for both variables. Sex was included as a covariate in all analyses, except for breast cancer, endometrial cancer, and female infertility, where only female controls were considered.

### Case series

We identified *MCM8/MCM9* variant carriers through multiple channels. On August 1, 2023, we conducted a comprehensive literature search for ‘MCM8’ and ‘MCM9’ in the NCBI PubMed database. This strategy yielded 116 studies discussing *MCM8* and 75 studies discussing *MCM9*. We included all studies in English and carefully examined them for any descriptions of *MCM8/MCM9* variant carriers. We excluded (systematic) reviews from to avoid duplicate patient data. Patient data was sourced from the papers themselves^22–32, 34–40, 45–50^ or updated data was obtained from the first or corresponding authors upon request.^2–4, 51^

Secondly, as part of the European Reference Network (ERN) for all patients with one of the rare genetic tumour risk syndromes (GENTURIS) initiative^52^, we identified *MCM8/MCM9* variant carriers not previously documented through outpatient clinics at various institutes across Europe. Patient data was sourced from genetic practitioners or retrieved from patient records.

#### Pathogenicity-based filtering and classification of the identified MCM8/MCM9 variant carriers

The *MCM8/MCM9* variants identified in the case series and cancer-specific cohorts (see next section) were filtered based on their predicted pathogenicity (**Figure 1**). Initially, we annotated the *MCM8/MCM9* variants using the guidelines from the American College of Medical Genetics and Genomics (ACMG) and the Association for Molecular Pathology (AMP) for variant interpretation^53, 54^, along with the CADD scoring^42^ and gnomAD v.2.1.1 allele frequency, accessed through Franklin.^55^ We excluded from the analysis (i) carriers of benign or likely benign *MCM8/MCM9* variants and (ii) variants of uncertain significance (VUS) with a CADD score lower than 20 or a gnomAD AF higher than 1%.

Individuals with homozygous or compound heterozygous variants that met the pathogenicity-based filtering criteria were considered as biallelic carriers. Conversely, compound heterozygous carriers with one variant meeting the criteria and one that did not were categorized as monoallelic carriers. Additionally, compound heterozygous carriers with a pathogenic or likely pathogenic variant and a VUS that met the pathogenicity-based filtering criteria were included in the pathogenic or likely pathogenic group, i.e., as biallelic carriers.

### Cancer-specific cohorts

We ascertained several cancer-specific cohorts to search for variant carriers with cancer phenotypes associated with germline *MCM8/MCM9* variants. These included 44 non-related serrated polyposis patients (SPS cohort)^56^ and 24 cancer-affected members of 16 nonpolyposis CRC families (fCRCX cohort) from which germline whole exome sequencing (WES) data was available for the analysis of single nucleotide variants and insertion-deletion mutations, as well as 632 metastasized CRCs and 25 metastasized endometrial carcinomas with available germline and tumor whole genome sequencing (WGS) data, accessible upon request by the Hartwig Medical Foundation database (reference number HMF-DR-288; https://www.hartwigmedicalfoundation.nl/). The identified *MCM8*/*MCM*9 variant carriers were filtered and classified based on pathogenicity using the same criteria applied to the *MCM8*/*MCM9* variant carriers from the case series, as detailed in the previous section (**Figure 1**).

### Mutational signature analysis

#### DNA sequencing and bioinformatic analysis of tumors from the case series

##### Patients and samples

To explore single base substitution (SBS) mutational signatures potentially associated with MCM8/MCM9 deficiency, DNA was obtained from formalin-fixed, paraffin-embedded (FFPE) tumor tissue from the following individuals of our case series: one individual (1 tumor) with biallelic *MCM8* variants, two individuals (5 tumors) with monoallelic *MCM8* variants, three individuals (10 tumors) with biallelic *MCM9* variants, and one individual (1 tumor) with monoallelic *MCM8* and *MCM9* variants.

##### Sample preparation and molecular evaluation

DNA extraction from FFPE tissue blocks was conducted using the NucleoSpin® DNA FFPE XS kit (MACHEREY-NAGEL, Düren, DE), and DNA concentrations were quantified using the Qubit™ Meter dsDNA High Sensitivity kit (ThermoFisher Scientific, Waltham, US). To ensure an adequate DNA level for subsequent analysis, DNA from a melanoma and an adenoma of one biallelic *MCM9* variant carrier (sample ID P5_34AB) were combined. WGS or WES was performed using the NovaSeq 6000 Sequencing System (Illumina Inc, San Diego, CA).

##### Somatic mutation calling

FASTQ files were aligned to the human genome build GRCh38.d1.vd1 using Burrows-Wheeler Aligner (BWA-MEM, v0.7.17).^57^ Picard MarkDuplicates (GATK v4.1.4.1) (Picard Toolkit, http://broadinstitute.github.io/picard; Broad Institute, Cambridge, US) was applied to mark all duplicated reads.^58^ SBS were identified using Mutect2 (GATK v4.1.4.1)^59^, VarScan (v2.4.3)^60^, MuSE (v1.0)^61^, and Strelka (v2.9.10)^62^ and filtered by variant caller confidences scores. Only variants that were called from at least two of these four callers were selected for the following mutational signature analysis and additional filtering based on their mutation confidence scores was applied: TLOD score >= 10 (Mutect2) and SomaticEVS >= 13 (Strelka2). Samples with no matched germline sequencing data (11 out of 16 samples) were applied only to Mutect2 for variant calling under tumor-only mode.

##### Mutational signature analysis

Mutational signature assignment was performed using SigProfilerAssignment (v0.0.32)^63^ based on the COSMICv3.3 SBS reference signatures.^64–68^ Treatment-associated signatures (SBS11, SBS25, SBS31, SBS32, SBS35, SBS86, SBS87, SBS90, and SBS99) were excluded from all samples before signature assignment (using the *exclude_signature_subgroups* option), except for sample ID P8_33A, who had history of neoadjuvant chemotherapy treatment.

#### Bioinformatic analysis of publicly available whole genome sequencing data

To further evaluate potential SBS mutational signatures associated with MCM8/MCM9 deficiency, we analyzed tumor WGS data from two publicly available sources. Firstly, we examined tumor data from cases with germline monoallelic *MCM8/MCM9* variants from the HMF cancer-specific cohort, as described earlier. Additionally, we evaluated tumor data from the TCGA Pan-Cancer Atlas, accessed through cBioPortal for Cancer Genomics (https://www.cbioportal.org/) between February and April of 2023.

For the TCGA Pan-Cancer Atlas samples, tumors from any cancer type were selected based on the presence of somatic *MCM8/MCM9* variant(s) that met the following criteria: (i) a tumor variant allele frequency of ≥ 20%; (ii) classified as pathogenic or likely pathogenic or as a VUS according to the ACMG/AMP recommendations for variant interpretation.^53, 54^ Additionally, variants were excluded in case they were assessed as tolerated by the Sorting Intolerant from Tolerant score^69^ and deemed benign by the PolyPhen score (**Figure 1**).^70^

In both the HMF and TCGA WGS datasets, SBS mutational signatures were identified by fitting the counts of single nucleotide variants (SNVs) per 96 tri-nucleotide context to the COSMIC v3.3 reference mutational signatures^71^, using the MutationalPatterns tool.^72^

### Statistical analysis

Clinical data was collected using Castor EDC (Castor Electronic Data Capture; https://castoredc.com). Figures were created, and statistical analysis was performed using Rstudio v2022.02.3+492 (Team R, Integrated Development for R, Boston, MA, 2022) or PLINK v1.9.

## Results

### Population-based cohorts

#### Individuals with (biallelic) germline MCM8/MCM9 variants are rare in gnomAD v.2.1.1

The occurrence of pLoF variants of *MCM8* in gnomAD (v.2.1.1) was 1.4 individuals per 100,000 persons across all populations, with the highest prevalence (5.5 per 100,000 persons) in the African/African American population (**Table 1**). Regarding *MCM9*, the prevalence of a pLoF variant was 2.5 individuals per 100,000 persons across all populations, with the highest prevalence (5.7 per 100,000 persons) found in the European Finnish population. The prevalence of missense *MCM8* and *MCM9* variants was 462.4 and 1173.3 individuals per 100,000 persons, respectively. Twenty-three individuals (0.02%) were identified as biallelic carriers of missense variants or more severe mutations of the *MCM8* gene (**Table 1**). With respect to the *MCM9* gene, 22 (0.02%) individuals were predicted to be biallelic carriers, including 21 carriers of missense variants or worse and one carrier of a homozygous pLoF variant.

**Table 1.**
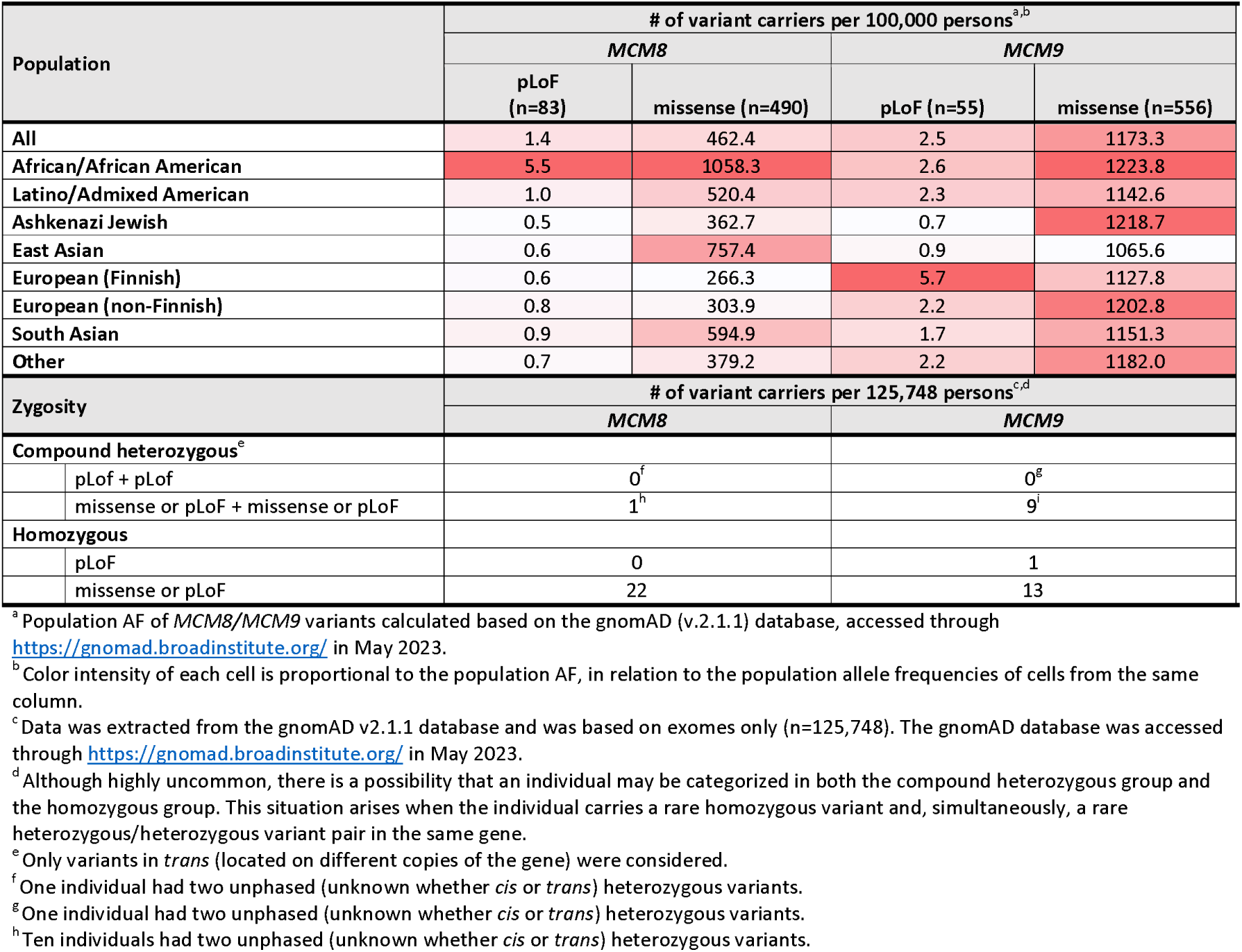
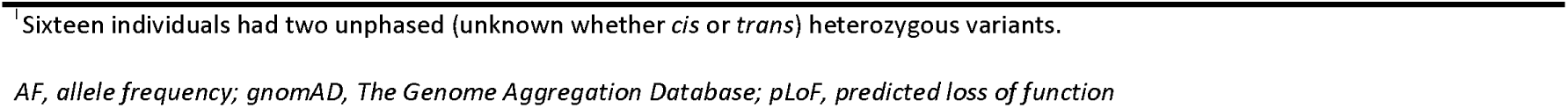
Population allele and biallelic carrier frequencies in gnomAD v.2.1.1.

| Population |  | # of variant carriers per 100,000 persons<sup>a,b</sup> |  |  |  |
| --- | --- | --- | --- | --- | --- |
|  |  | MCM8 |  | MCM9 |  |
|  |  | pLoF (n=83) | missense (n=490) | pLoF (n=55) | missense (n=556) |
| All |  | 1.4 | 462.4 | 2.5 | 1173.3 |
| African/African American |  | 5.5 | 1058.3 | 2.6 | 1223.8 |
| Latino/Admixed American |  | 1.0 | 520.4 | 2.3 | 1142.6 |
| Ashkenazi Jewish |  | 0.5 | 362.7 | 0.7 | 1218.7 |
| East Asian |  | 0.6 | 757.4 | 0.9 | 1065.6 |
| European (Finnish) |  | 0.6 | 266.3 | 5.7 | 1127.8 |
| European (non-Finnish) |  | 0.8 | 303.9 | 2.2 | 1202.8 |
| South Asian |  | 0.9 | 594.9 | 1.7 | 1151.3 |
| Other |  | 0.7 | 379.2 | 2.2 | 1182.0 |
| Zygosity |  | # of variant carriers per 125,748 persons<sup>c,d</sup> |  |  |  |
|  |  | MCM8 |  | MCM9 |  |
| Compound heterozygous<sup>e</sup> |  |  |  |  |  |
|  | pLoF + pLoF | 0<sup>f</sup> |  | 0<sup>g</sup> |  |
|  | missense or pLoF + missense or pLoF | 1<sup>h</sup> |  | 9<sup>i</sup> |  |
| Homozygous |  |  |  |  |  |
|  | pLoF | 0 |  | 1 |  |
|  | missense or pLoF | 22 |  | 13 |  |

<sup>a</sup> Population AF of *MCM8/MCM9* variants calculated based on the gnomAD (v.2.1.1) database, accessed through
https://gnomad.broadinstitute.org/ in May 2023.
<sup>b</sup> Color intensity of each cell is proportional to the population AF, in relation to the population allele frequencies of cells from the same column.
<sup>c</sup> Data was extracted from the gnomAD v2.1.1 database and was based on exomes only (n=125,748). The gnomAD database was accessed through https://gnomad.broadinstitute.org/ in May 2023.
<sup>d</sup> Although highly uncommon, there is a possibility that an individual may be categorized in both the compound heterozygous group and the homozygous group. This situation arises when the individual carries a rare homozygous variant and, simultaneously, a rare heterozygous/heterozygous variant pair in the same gene.
<sup>e</sup> Only variants in *trans* (located on different copies of the gene) were considered.
<sup>f</sup> One individual had two unphased (unknown whether *cis* or *trans*) heterozygous variants.
<sup>g</sup> One individual had two unphased (unknown whether *cis* or *trans*) heterozygous variants.
<sup>h</sup> Ten individuals had two unphased (unknown whether *cis* or *trans*) heterozygous variants.
<sup>1</sup>Sixteen individuals had two unphased (unknown whether *cis* or *trans*) heterozygous variants.

#### Biallelic MCM9 variant carriers in the 100K Genomes Project have an increased risk of polyposis and gastric cancer, while no enrichment was observed for biallelic MCM8 variants or in the 200K UK Biobank dataset

In the 100K Genomes Project, we identified 51 biallelic carriers (21 homozygous and 30 compound heterozygous) and 2,782 monoallelic carriers of pLoF or predicted deleterious missense variants in the *MCM8* gene. Moreover, we found 64 biallelic carriers (21 homozygous and 43 compound heterozygous) and 3,166 monoallelic carriers of pLoF or predicted deleterious missense variants in the *MCM9* gene. Among the 51 biallelic *MCM8* variant carriers in the 100K Genomes Project, 2 individuals (3.9%) had colorectal cancer (CRC), 3 (5.9%) had colonic polyps, 3 (5.9%) had colonic adenomas, 3 (5.9%) had rectal polyps, 2 (3.9%) had hypothyroidism, and 5 (9.8%) had breast cancer. Additionally, 1 individual (2.0%) had epilepsy, 1 had endometrial cancer, 1 had short stature, and 1 experienced delayed puberty. Among the 64 biallelic *MCM9* variant carriers in the 100K Genomes Project, 3 individuals (4.7%) had colorectal cancer (CRC), 2 (3.1%) had colonic polyps, 2 (3.1%) had colonic adenomas, 2 (3.1%) had rectal polyps, 3 (4.7%) had hypothyroidism, 5 (7.8%) had breast cancer, and 2 (3.1%) had epilepsy. Additionally, 1 individual (1.6%) had melanoma, 1 had gastric cancer, and 1 had endometrial cancer. While no significant enrichment of biallelic *MCM8* pLoF or predicted deleterious missense variants was observed for any of these phenotypes compared to controls, we did observe significant associations between biallelic *MCM9* pLoF or predicted deleterious missense variants and colonic polyps (odds ratio (OR) 6.51, 95% confidence interval (CI) 1.24-34.11; *P* = 0.03), rectal polyps (OR 8.40, 95% CI 1.28-55.35; *P* = 0.03), and gastric cancer (OR 27.03, 95% CI 2.93-248.5; *P* = 0.004) (**Table 2**).

**Table 2.**
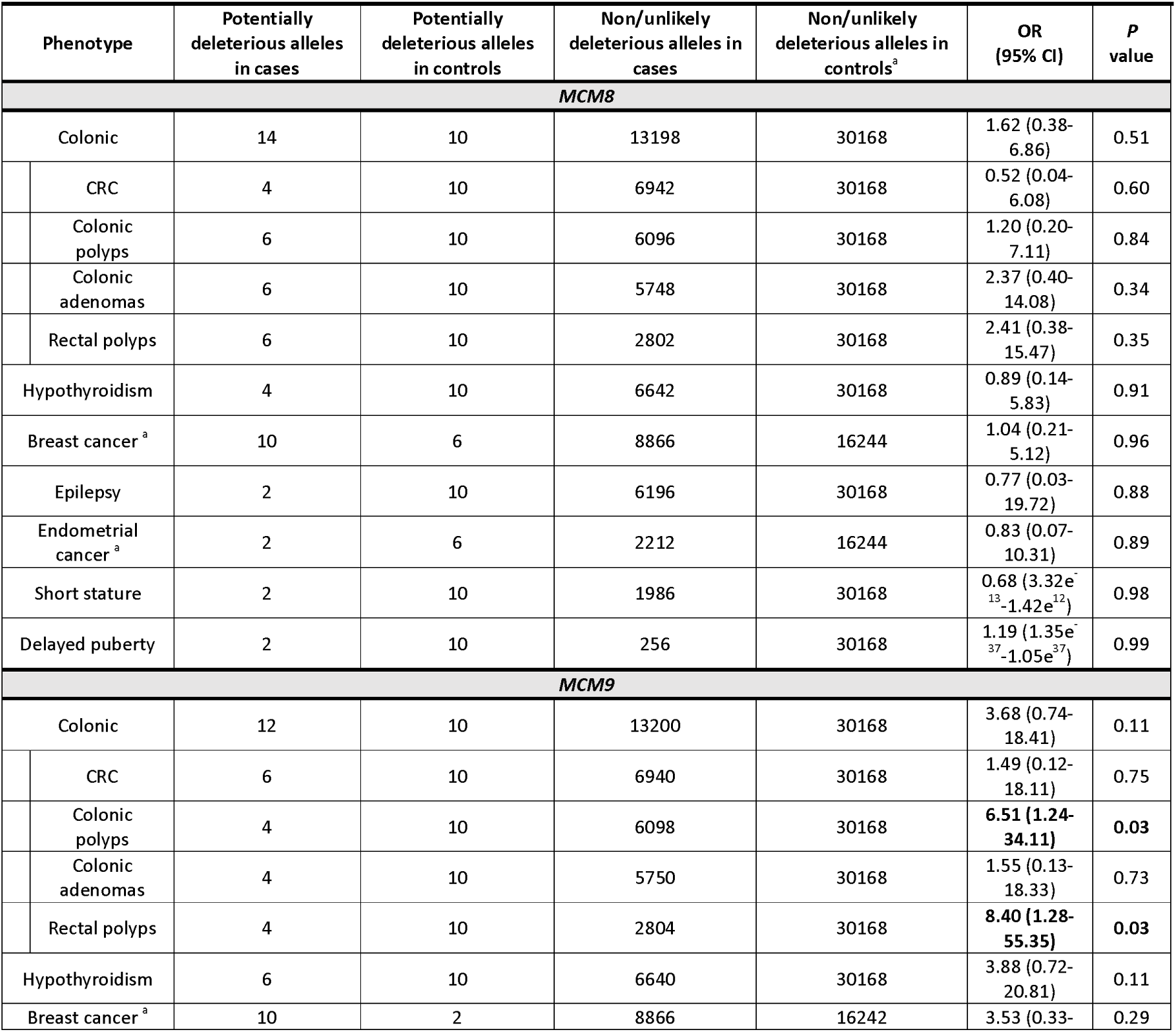
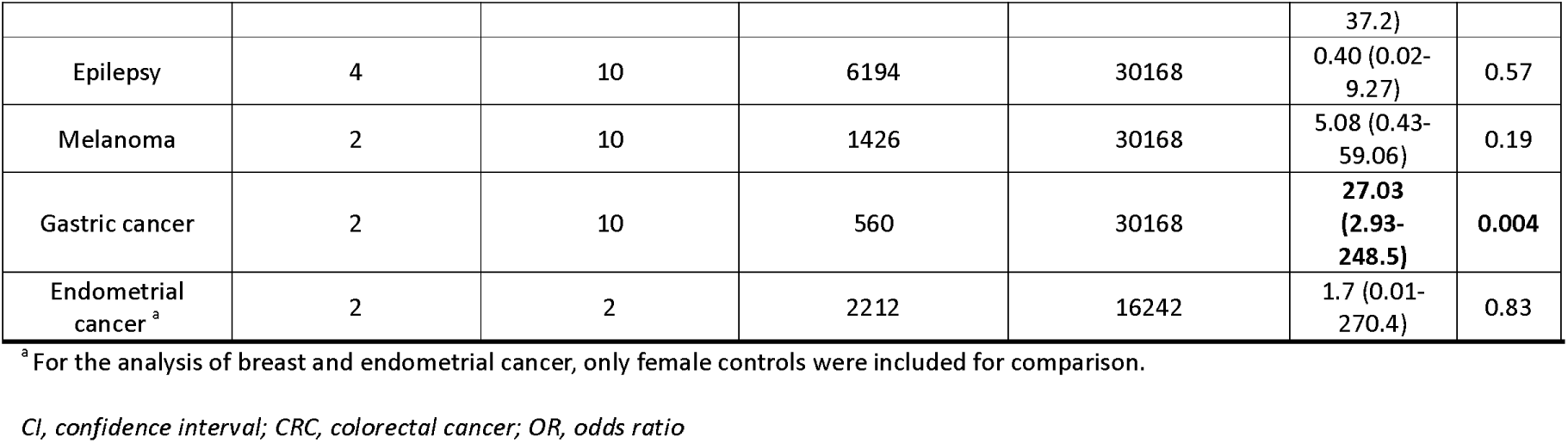
Enrichment analysis of biallelic MCM8/MCM9 variants in 100K Genomes Project, adjusting for age, sex and ethnicity.

In the 200K exomes release of the UK Biobank, we identified 110 biallelic carriers (47 homozygous and 63 compound heterozygous) and 8,453 monoallelic carriers of pLoF or predicted deleterious missense variants in the *MCM8* gene. Additionally, we found 74 biallelic carriers (15 homozygous and 59 compound heterozygous) and 4,991 monoallelic carriers of pLoF or predicted deleterious missense variants in the *MCM9* gene. Among the 110 biallelic *MCM8* variant carriers in the 200K UK Biobank, 2 individuals (1.8%) were registered with CRC, 3 (2.7%) with colonic polyps, 4 (3.6%) with adenomas, 1 (0.9%) with female infertility, and 6 (5.5%) with hypothyroidism. Among the 74 biallelic *MCM9* variant carriers in the 200K UK Biobank, 1 individual (1.4%) was registered with colorectal cancer (CRC), 3 (4%) with colonic polyps, 6 (8%) with adenomas, 1 (1.4%) with rectal polyps, and 2 (2.7%) with hypothyroidism. However, no significant enrichment of biallelic *MCM8*/*MCM9* pLoF or predicted deleterious missense variants was observed for any of these phenotypes compared to controls in the 200K UK Biobank (**Table 3**).

**Table 3.** Enrichment analysis of biallelic MCM8/MCM9 variants in 200K UK Biobank, adjusting for age, sex and ethnicity.

| Phenotype | Potentially deleterious alleles in cases | Potentially deleterious alleles in controls | Non/unlikely deleterious alleles in cases | Non/unlikely deleterious alleles in controls<sup>a</sup> | OR (95% CI) | P value |
| --- | --- | --- | --- | --- | --- | --- |
| ***MCM8*** |  |  |  |  |  |  |
| Colonic | 16 | 136 | 39158 | 181658 | 0.54 (0.26-1.14) | 0.11 |
| CRC | 4 | 136 | 6474 | 181658 | 0.83 (0.20-3.41) | 0.80 |
| Colonic polyps | 6 | 136 | 18518 | 181658 | 0.43 (0.14-1.39) | 0.16 |
| Colonic adenomas | 8 | 136 | 20586 | 181658 | 0.52 (0.19-1.44) | 0.21 |
| Rectal polyps | 0 | 136 | 10328 | 181658 | NA | NA |
| Female infertility<sup>a</sup> | 2 | 74 | 988 | 98908 | 2.68 (0.34-21.11) | 0.35 |
| Hypothyroidism | 12 | 136 | 21982 | 181658 | 0.67 (0.29-1.58) | 0.36 |
| ***MCM9*** |  |  |  |  |  |  |
| Colonic | 16 | 84 | 39158 | 181710 |  |  |
| CRC | 2 | 84 | 6476 | 181710 | 0.82 (0.11-5.99) | 0.84 |
| Colonic polyps | 6 | 84 | 18518 | 181710 | 0.80 (0.25-2.63) | 0.72 |
| Colonic adenomas | 12 | 84 | 20582 | 181710 | 1.51 (0.63-3.61) | 0.35 |
| Rectal polyps | 2 | 84 | 10326 | 181710 | 0.49 (0.07-3.59) | 0.48 |
| Female infertility<sup>a</sup> | 0 | 42 | 990 | 98898 | NA | NA |
| Hypothyroidism | 4 | 84 | 21990 | 181710 | 0.46 (0.11-1.94) | 0.29 |

<sup>a</sup> For the analysis of female infertility, only female controls were included for comparison.
CI, confidence interval; CRC, colorectal cancer; NA, not applicable; OR, odds ratio

None of the other phenotypes investigated (see **Supplementary Table 1**) were registered among the biallelic *MCM8*/*MCM9* variant carriers, based on ICD10/ICD-O registrations.

### Case series

#### Phenotype of biallelic germline MCM8/MCM9 variants carriers

In our case series, we identified 26 biallelic *MCM8* variant carriers (including 15 with pathogenic or likely pathogenic variants and 11 with a VUS) and 28 biallelic *MCM9* variant carriers (including 22 with pathogenic or likely pathogenic variants and 6 with a VUS) that met the pathogenicity-based filtering criteria. This group included 3 biallelic *MCM8* and 4 biallelic *MCM9* variant carriers who had not been previously described (**Figure 1**). An overview of all identified *MCM8/MCM9* variant carriers, including their sources, is presented in **Supplementary Table 2**. A detailed description of all newly identified *MCM8/MCM9* variant carriers and previously documented carriers for whom we obtained updated clinical information (individuals meeting the pathogenicity-based filtering criteria only), including pedigrees, is available from the corresponding author upon reasonable request.

#### Biallelic *MCM8*/*MCM9* variant carriers often present with hypogonadism linked to impaired gonadal development

The majority of individuals with biallelic *MCM8* (23 out of 26, 88%) or *MCM9* (26 out of 28, 93%) variants from our case series experienced hypogonadism (<u>HP:0000815</u>) (**Figure 2**). Apart from five males (three with biallelic *MCM8* variants and two with biallelic *MCM9* variants) with azoospermia (no sperm in the semen; <u>HP:0000027</u>), these issues involved women affected by POI. Fourteen out of twenty (70%) individuals affected by POI and carrying biallelic *MCM8* variants had undetectable or small ovaries coupled with an infantile or absent uterus upon ultrasound in thirteen (65%) of the cases. Among the biallelic *MCM9* variant carriers affected by POI, 14 out of 23 (61%) exhibited invisible or small ovaries, and 12 out of 23 (52%) had infantile or absent uteri. Furthermore, osteoporosis or delayed bone age (<u>HP:0000939</u>) was reported in seven individuals with biallelic *MCM9* variants and one individual with biallelic *MCM8* variants, which were all affected by hypogonadism. In both the *MCM8* and *MCM9* groups, hypogonadism manifested at a relatively young age, typically between 10 and 30 years (**Figure 3**). Many of these patients were part of earlier studies, with no updated clinical data available upon request, so most were lost to follow-up post-publication.

**Figure 2.**
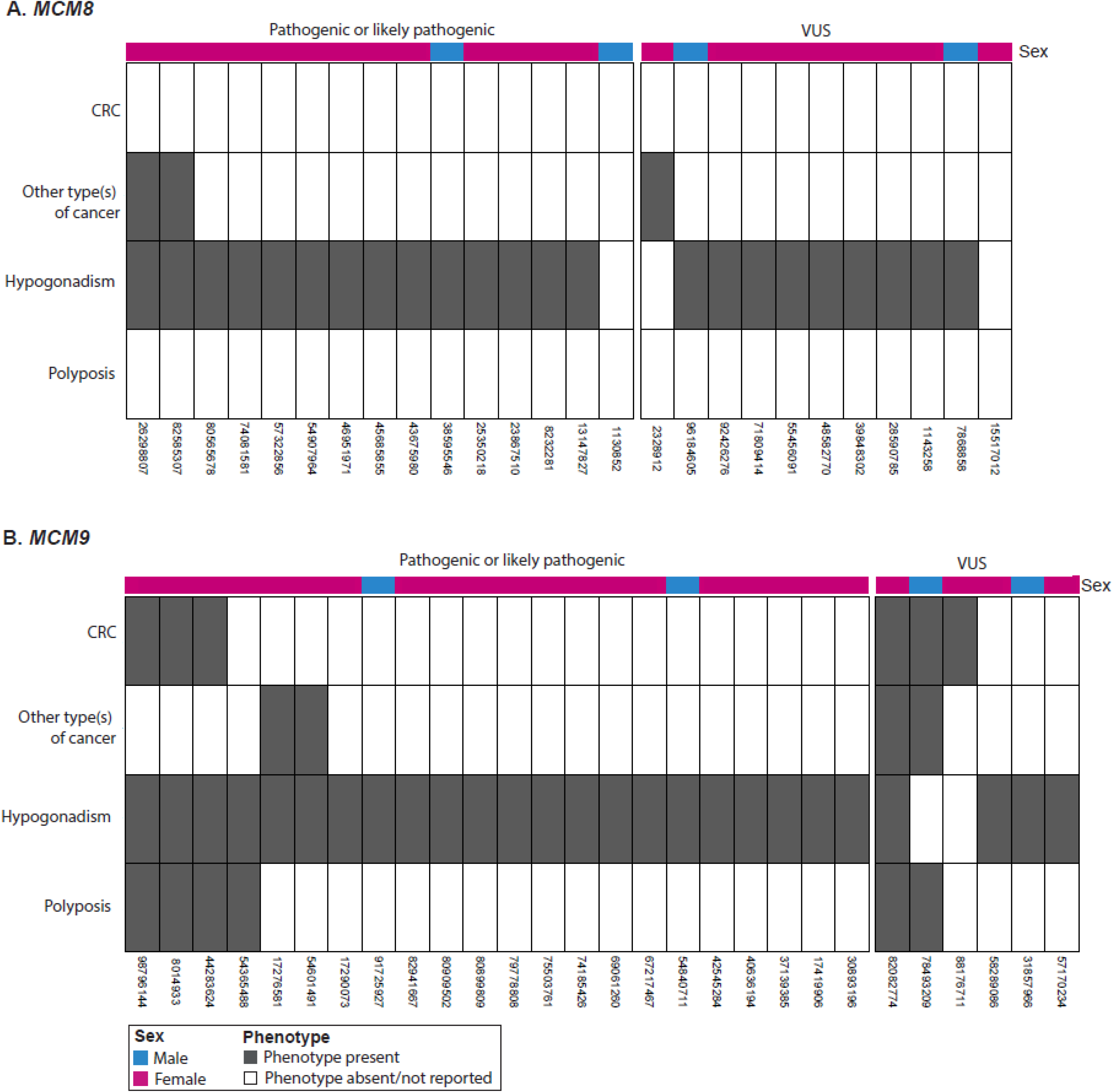
Phenotype of biallelic *MCM8/MCM9* variant carriers. The phenotype is presented for all (**A**) biallelic *MCM8* and (**B**) biallelic *MCM9* variant carriers from our case series. Each column represents an individual, while each row corresponds to one of the four primary observed phenotypes: CRC, other type(s) of cancer, hypogonadism, and polyposis. Person IDs are provided below each column. *CRC, colorectal cancer; VUS, variant of uncertain significance*.

**Figure 3.**
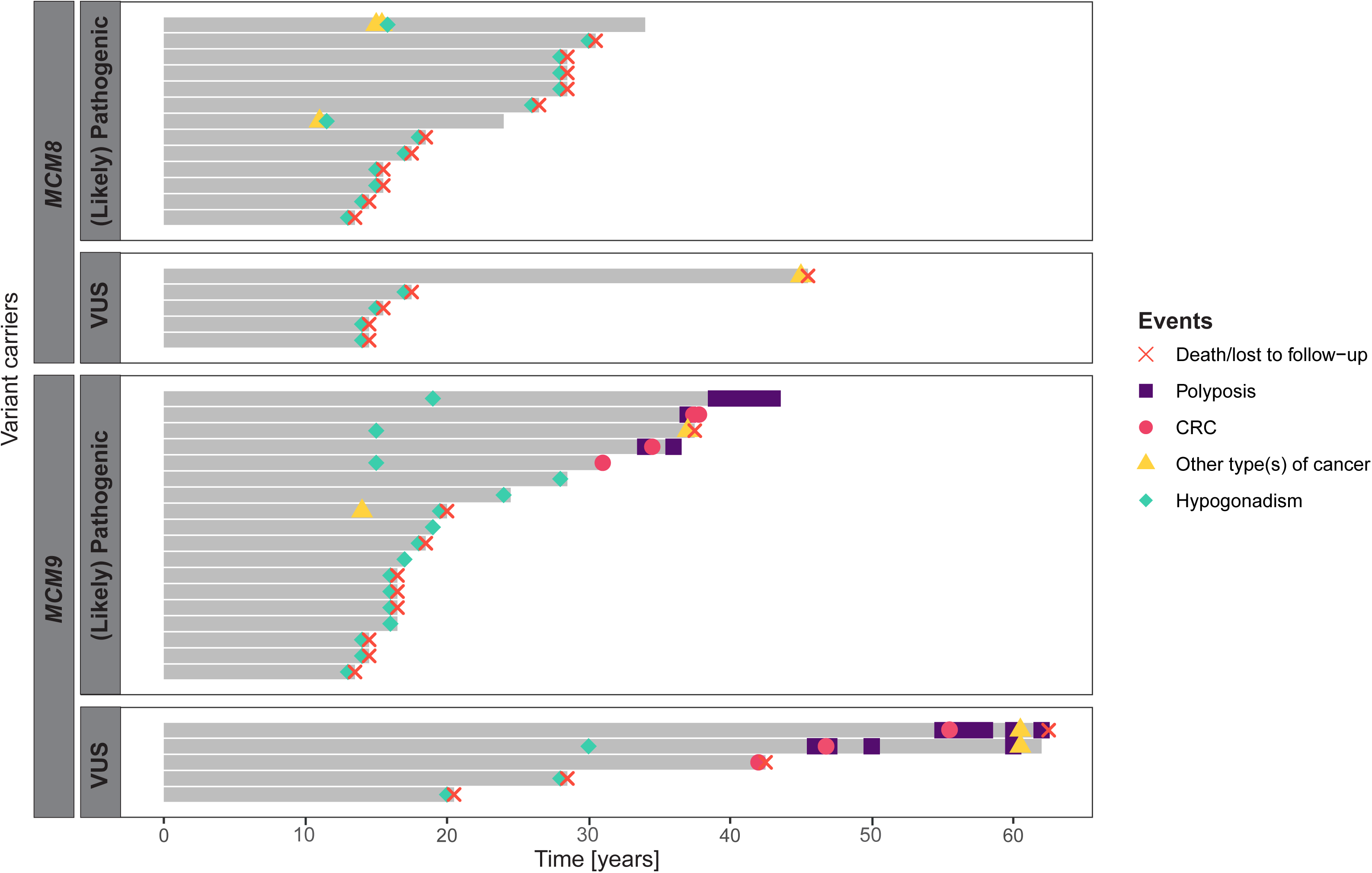
Disease onset in biallelic *MCM8/MCM9* variant carriers. The onset of the four primary observed phenotypes (CRC, other type(s) of cancer, hypogonadism, and polyposis) is displayed for each biallelic *MCM8/MCM9* variant carrier with available age details in our case series. Those without age details were excluded from the analysis. Individuals are ordered by ACMG/AMP classification (pathogenic or likely pathogenic, VUS)^53, 54^ and current age or age at the time of death/lost to follow-up. *ACMG, American College of Medical Genetics and Genomics; AMP Association for Molecular Pathology; CRC, colorectal cancer; VUS, variant of uncertain significance*.

#### Biallelic *MCM9* variant carriers may face polyposis, gastric cancer and early-onset colorectal cancer, while both biallelic *MCM8*/*MCM9* carriers may face female germ cell tumors

Polyposis (typically > 20 polyps, including hyperplastic, adenomatous, and serrated types) was reported in 6 out of 28 (21%) biallelic *MCM9* variant carriers from our case series (**Figure 2**). Similarly, CRC was observed in 6 of 28 (21%) biallelic *MCM9* variant carriers in our case series. This includes three carriers of likely pathogenic variant(s) who developed CRC between the ages of 30 and 40, and three carriers with a variant of uncertain significance (VUS) diagnosed between 40 and 60 years (**Figure 3**). No CRC or polyp diagnoses were reported among the biallelic *MCM8* variant carriers. Three female carriers—two with biallelic *MCM8* variants and one with a biallelic *MCM9* variant—were diagnosed with germ cell tumors (<u>HP:0100728</u>) between the ages of 11 and 15 years. These included two endodermal sinus tumors originating from dysgerminomas, which themselves arose from gonadoblastomas, and one germ cell tumor of unspecified origin. Single biallelic *MCM9* variant carriers were diagnosed with gastric cancer (<u>HP:0012126</u>), an HPV-unrelated clear cell carcinoma of the cervix (<u>HP:0031522</u>), and melanoma (<u>HP:0012056</u>), whereas a biallelic *MCM8* variant carrier was diagnosed with breast cancer (<u>HP:0003002</u>).

#### Monoallelic *MCM8*/*MCM9* variants may experience hypogonadism

During the pathogenicity-based filtering process of our case series, we filtered 49 mono-allelic *MCM8* variant carriers and 45 monoallelic *MCM9* variant carriers. Out of these 49 monoallelic *MCM8* variant carriers, hypogonadism was noted in 14 (29%) individuals, with two having a likely pathogenic variant and 12 carrying a VUS (**Supplementary Figure 1-2**). Two monoallelic *MCM8* variant carriers were diagnosed with CRC, another two with polyposis, and two individuals with a monoallelic *MCM8* variant were diagnosed with breast cancer.

Among the 45 monoallelic *MCM9* variant carriers from our case series, ten (22%) were known to have hypogonadism, including one individual who was also diagnosed with CRC and polyposis (**Supplementary Figure 1-2**). CRC and polyps were additionally reported in five and six other monoallelic *MCM9* variant carriers, respectively. No other types of cancer were reported in the monoallelic *MCM9* group.

#### Genotype-phenotype correlations reveal potential hotspot sites

Mapping of variants onto the MCM8 and MCM9 protein domains revealed that the variants in our case series clustered in two key regions: the N-terminal DNA binding domain, which is crucial for protein-DNA binding (6 of 11 *MCM8* variants in biallelic carriers, 55%; 4 of 20 *MCM9* variants in biallelic carriers, 20%), and the AAA+ core domain, essential for DNA helicase activity (5 of 11 *MCM8* variants in biallelic carriers, 45%; 12 of 20 *MCM9* variants in biallelic carriers, 60%) (**Figure 4**, **Supplementary Figure 3**).^44^ Additionally, several variants were found to be shared among multiple families with hypogonadism from our case series. For instance, the c.482A>C [p.(His161Pro)] VUS in the *MCM8* gene, previously linked to hypogonadism^26, 39, 46, 49, 51^, was shared by six biallelic carriers across two families. Similarly, the pathogenic c.394C>T [p.(Arg132*)] variant in the *MCM9* gene, also associated with hypogonadism^39, 49, 51^, was shared by seven biallelic carriers from four unrelated families.

**Figure 4.**
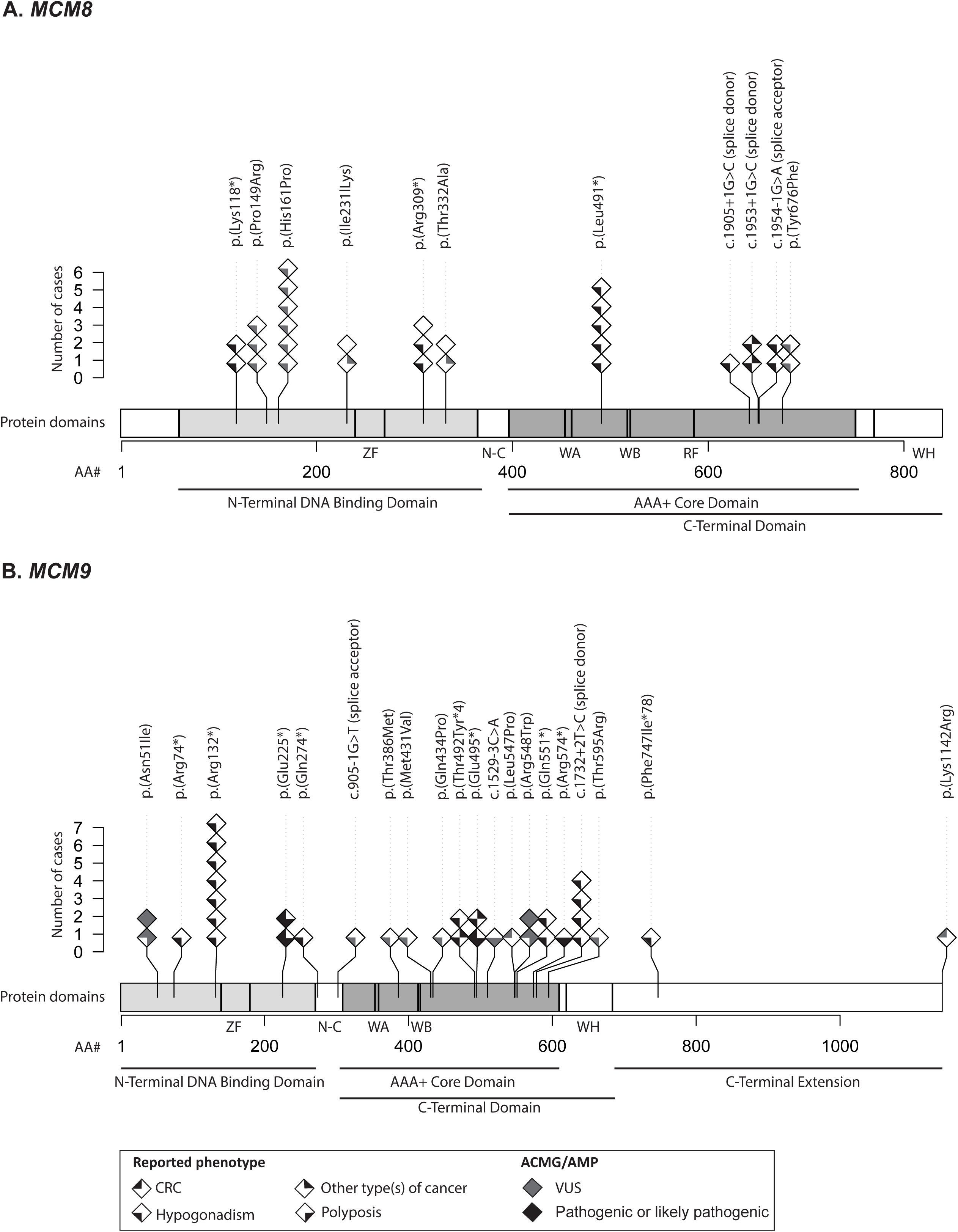
Biallelic *MCM8/MCM9* variants mapped onto the respective protein domains. (**A**) *MCM8* and (**B**) *MCM9* variants from all biallelic variant carriers in our case series are mapped onto the domains of the MCM8 and MCM9 proteins, respectively. Each homozygote variant carrier corresponds to one diamond symbol, whereas for compound heterozygous variant carriers, both variants are separately plotted. The fill and color of the diamond symbols correspond to the phenotype of the individual (CRC, other type(s) of cancer, hypogonadism, polyposis) and the ACMG/AMP classification of the variant (pathogenic or likely pathogenic, VUS)^53, 54^, respectively. *ACMG, American College of Medical Genetics and Genomics; AMP, Association for Molecular Pathology; CRC, colorectal cancer; RF, arginine finger; VUS, variant of uncertain significance; WA, Walker A; WB, Walker B; WH, winged-helix; ZF, zinc-finger*.

### Cancer-specific cohorts

No biallelic *MCM8/MCM9* variant carriers meeting the pathogenicity-based filtering criteria were identified in the SPS (serrated polyposis cases), fCRCX (nonpolyposis CRC families) and HMF (metastasized CRC and endometrial cancer cases) cancer-specific cohorts. In the HMF cancer-specific cohort, four monoallelic *MCM8* and three monoallelic *MCM9* variant carriers meeting the pathogenicity-based filtering criteria were identified with CRC.

### Mutational signature analysis

#### Clock-like mutational processes dominate in tumors from the case series and HMF cancer-specific cohort

SBS mutational signatures SBS1 and SBS5, which represent clock-like mutational processes^73^, were present across all tumors from our case series with matched germline sequencing data available (5 out of 16 sequenced tumors, 31.3%, involving three CRCs and two breast cancers), and were similarly present in a CRC and two polyps from a wildtype control (**Figure 5A-B**). With respect to MMR deficiency-associated signatures, only signature SBS26 was detected and only in one tumor (P2_2T, breast cancer), where it contributed to a minority of the mutations (269 out of 2026, 13%) (**Figure 5B**).^64–68^ Sequenced tumors without matched germline sequencing data available were dominated by sequencing artefact signatures (SBS45, SBS47, SBS50, SBS51, SBS54, SBS56, SBS58, SBS95)^64–68^, precluding us from the possibility of comparing these cases to those with matched controls or previously published cases.^4^ Clock-like signatures SBS1 and SBS5, along with signatures SBS93 and SBS40 of unknown etiology, were the most prevalent signatures in metastasized CRCs from seven individuals carrying monoallelic *MCM8*/*MCM9* variants in the HMF cancer-specific cohort (**Supplementary Figure 4**).

**Figure 5.**
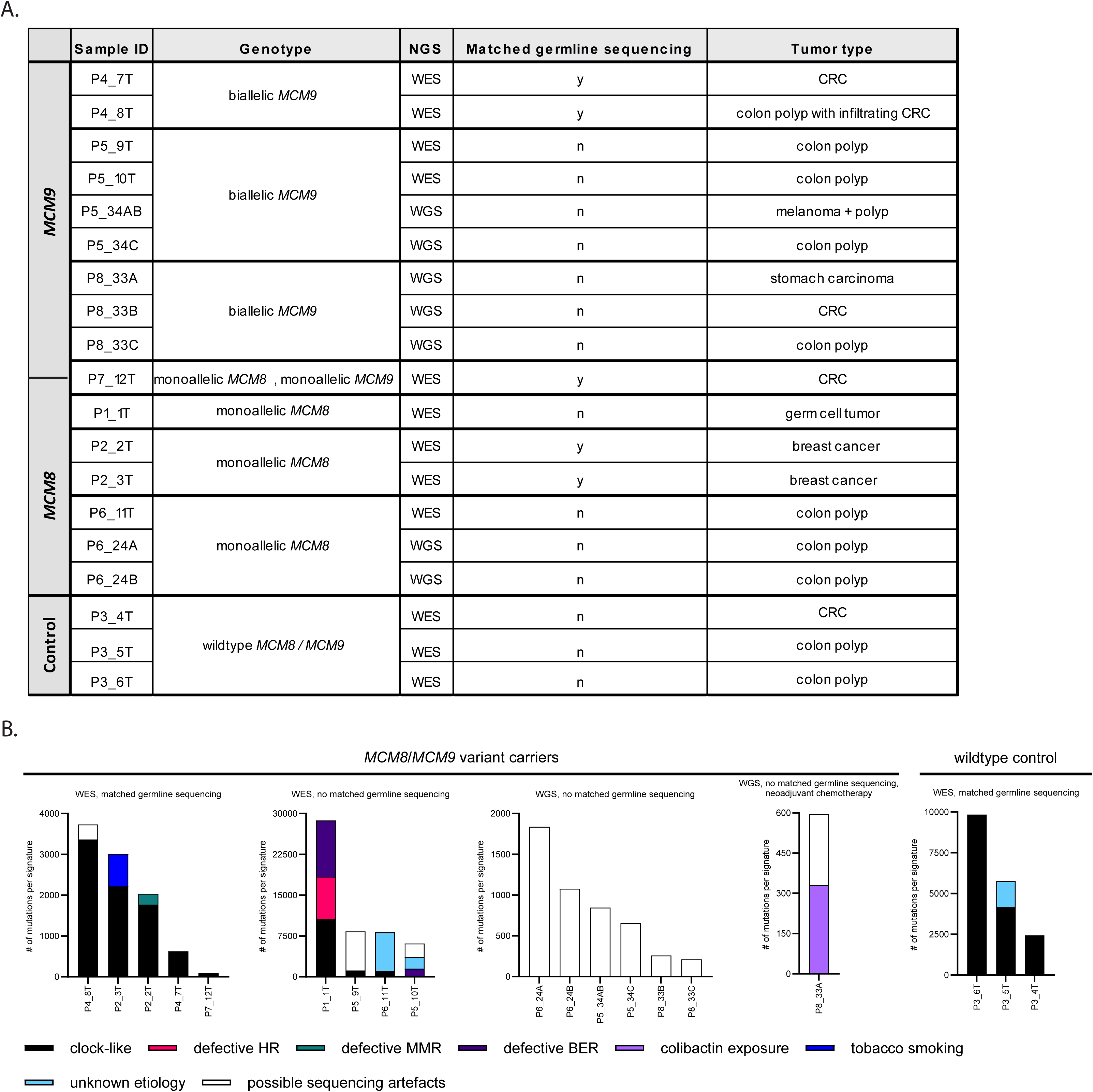
Mutational signatures analysis of tumors from *MCM8/MCM9* variant carriers from our case series. (**A**) Sample overview of the tumors that were available from our case series for mutational signature analysis, including the corresponding genotype, next generation sequencing (NGS) approach, the availability of normal control tissue, and the tumor type. Control tissue originated from an individual tested negative for germline *MCM8/MCM9* variants. (**B**) The number of mutations in each signature is presented for every tumor. Mutational signature assignment was performed using SigProfilerAssignment (v0.0.32)^63^ based on the COSMICv3.3 single base substitution (SBS) reference signatures. SBS1 and SBS5 were classified as clock-like mutational signatures, while SBS3 was considered to be caused by defective homologous recombination (HR) repair. SBS26 was linked to defective MMR, and SBS30 and SBS36 were associated with defective base excision repair (BER). SBS88 was attributed to colibactin exposure, and SBS92 to tobacco smoking. SBS37, SBS40, and SBS94 were considered to be of unknown etiology, while SBS40, SBS45, SBS50, SBS51, SBS54, SBS56, SBS58, and SBS95 were considered possible sequencing artifacts. *BER, base excision repair; CRC, colorectal cancer; HR, homologous repair; MMR, mismatch repair; NGS, next generation sequencing; SBS, single base substitution; WES, whole exome sequencing; WGS, whole genome sequencing*.

#### Somatic MCM8/MCM9 mutations may occur as a result of other DNA repair deficiencies and mutational processes, potentially involving copy number variations

In the TCGA Pan-Cancer Atlas dataset, insights into the somatic mutational behavior of *MCM8* and *MCM9* were gained through the observation of copy number alterations in both genes. Furthermore, unsupervised hierarchical clustering of SBS mutational signature profiles revealed clusters characterized by signatures such as SBS7a/b (UV damage), SBS2 and SBS13 (APOBEC activity), SBS6, SBS14, SBS15, SBS20, and SBS21 (MMR deficiency), and SBS10a/b (POLE deficiency)^64–68^, which suggest that somatic *MCM8*/*MCM9* variants may be secondary to other DNA repair deficiencies and mutational processes (**Supplementary Figure 5**).

## Discussion

Following the initial discovery of biallelic germline *MCM8*/*MCM9* variants in families with CRC, polyposis, and hypogonadism^2–4^, we present a comprehensive clinical and molecular characterization of biallelic *MCM8*/*MCM9* variant carriers from multiple sources. Our analysis of the 100K Genomes Project reveals that biallelic *MCM9* variant carriers are at increased risk for polyposis and gastric cancer, a pattern not observed in biallelic *MCM8* carriers. This finding is further supported by our case series, which included 26 biallelic *MCM8* and 28 biallelic *MCM9* variant carriers, including seven previously unreported cases. Furthermore, the case series indicates that, in addition to hypogonadism associated with impaired gonadal development, biallelic *MCM8* and *MCM9* variants are linked to the development of germ cell tumors, with biallelic *MCM9* variants potentially associated with early-onset CRC. These findings highlight the importance of including *MCM8* and *MCM9* in diagnostic gene panels for relevant clinical contexts and suggest that biallelic carriers may benefit from cancer surveillance.

Gaining an unbiased understanding of the phenotype of biallelic *MCM8/MCM9* variant carriers is currently challenging. This difficulty arises mainly from the limited inclusion of *MCM8/MCM9* genes in current diagnostic gene panels for cancer and polyposis, constraining our case series, and the relative rarity of germline *MCM8/MCM9* variants in the general population, as reflected by our investigations in gnomAD v.2.1.1, the 100K Genomes Project, and the 200K exome release of the UK Biobank. This rarity may have contributed to the absence of biallelic *MCM8*/*MCM9* variants in the cancer-specific cohorts and could have influenced the enrichment analysis of these variants in the 100K Genomes Project and 200K UK Biobank. Aside from the increased risk of polyposis and gastric cancer associated with biallelic *MCM9* variants in the 100K Genomes Project, the lack of enrichment for biallelic *MCM8*/*MCM9* variants in other phenotypes and in the 200K UK Biobank may be attributed to one of two factors: (i) these variants may not actually contribute to studied phenotypes, or (ii) there may be limitations in the analysis itself, such as reliance on the accuracy and consistency of ICD10/ICD-O registrations and the variant filtering approach, which, partly due to the relative novelty of both genes, relied primarily on *in silico* prediction tools. With regards to our case series, we acknowledge an ascertainment bias, contributing to the high frequency of hypogonadism in our cohort since most patients examined were from studies primarily focused on fertility problems rather than cancer. In contrast, the occurrence of cancer and polyposis among biallelic *MCM8/MCM9* variant carriers may be underestimated because many patients in our case series are still young, potentially too young to have developed cancer, and because colonoscopies are not typically recommended for biallelic *MCM8/MCM9* variant carriers. Moreover, the prevalence of the associated phenotypes might be underestimated due to our variant filtering approach, being dependent on limited *in silico* prediction algorithms and data from previous studies, for instance with regards to segregation analysis and variant phasing. This may have led to misclassification of individuals as (biallelic) variant carriers, thereby potentially diluting the observed prevalence of phenotypes in our analyses.

Despite its limitations, our population-based analysis and case series describe the most extensive collection of individuals with biallelic *MCM8*/*MCM9* variants to date, underscoring the importance of considering these variants in specific clinical contexts. We recommend considering biallelic *MCM9* variants in individuals and families with unexplained polyposis, gastric cancer, germ cell tumors, or (early-onset) CRC, particularly in cases of recessive inheritance and known hypogonadism, until more data are available. Similarly, biallelic *MCM8* variants should be considered in cases of unexplained germ cell tumors, especially when accompanied by recessive inheritance or hypogonadism. Additionally, given previous reports linking biallelic *MCM8* variants to CRC^4^ and the potential underestimation of cancer and polyposis in our case series, it may be prudent to consider biallelic *MCM8* variants in cases of unexplained CRC or polyposis until further data are available. As these genes become more integrated into diagnostic gene panels and more families are identified, larger sample sizes and longer follow-up periods will allow for more accurate cancer risk assessments.

Given the range of malignancies observed in our case series, surveillance for these patients could be considered within a shared decision-making framework, taking into account the current evidence until more data become available. Similar to the *NTHL1*-and *MUTYH*-deficiency syndromes^74–76^, which are associated with CRC and polyposis, the *MCM9*-deficiency syndrome observed in our population-based analysis and case series may warrant comparable surveillance protocols. Established colon surveillance guidelines for *NTHL1*-and *MUTYH*-deficiency syndromes^74–76^, which recommend (bi)annual colonoscopy beginning around 18-20 years of age, could potentially be extended to individuals carrying biallelic *MCM9* variants. However, given the observed onset age of 30–60 years in our series, initiating colonoscopy at 25 years may be more appropriate. Additionally, due to the potential increased risk of gastric cancer, concurrent gastroscopy could be considered. Considering the prevalence of germ cell tumors in female biallelic *MCM8/MCM9* variant carriers, annual ultrasound screening starting at age 10 could be considered, given the early onset of 11-15 years observed in our case series. Further evaluation of cancer risks and the cost-effectiveness of surveillance measures is necessary to develop comprehensive surveillance guidelines.

In contrast to biallelic *MCM8/MCM9* variant carriers, our current data suggest that the phenotype of monoallelic *MCM8*/*MCM9* variant carriers may primarily be limited to hypogonadism, with no clear evidence of an increased cancer risk, which does not seem to justify cancer surveillance for these individuals. Although the prevalence of hypogonadism among monoallelic carriers in our case series (29% for *MCM8*, 22% for *MCM9*) appears higher than the global prevalence (e.g., 3.5% for POI^77^), the potential ascertainment bias in our study, as previously discussed, highlights the need for further research to more fully characterize the phenotype of monoallelic *MCM8*/*MCM9* variant carriers.

To gain potential causal evidence for a role of MCM8/MCM9 deficiency in the development of polyps and cancer, future studies exploring the mutational signatures of tumors from *MCM8/MCM9* variant carriers are essential. In the mutational signature analysis from our case series, we observed that clock-like mutational processes dominate in tumors from *MCM8/MCM9* variant carriers with matched germline sequencing data available. However, we could not confirm the proposed role of MCM8 and MCM9 in MMR or other DNA repair mechanisms based on specific SBS signature contributions. The clock-like mutational signatures SBS1 and SBS5, commonly found in most CRCs without specific DNA repair defects and in many other cancer types^64–68^, were not more prevalent in tumors from *MCM8*/*MCM9* variant carriers than in those from our wildtype control. Future studies should therefore confirm whether tumors from *MCM8*/*MCM9* variant carriers are molecularly similar to sporadic tumors or if other mutational signatures, possibly missed in this study, are associated with MCM8/MCM9 deficiency.

In conclusion, our study offers a detailed clinical and molecular characterization of biallelic *MCM8*/*MCM9* variant carriers from various sources. Our data suggests that biallelic *MCM9* variants are associated with polyposis, gastric cancer, and early-onset CRC, while both biallelic *MCM8* and *MCM9* variants are linked to hypogonadism and the early development of germ cell tumors. These findings support the inclusion of *MCM8*/*MCM9* in diagnostic gene panels for specific clinical contexts and indicate that carriers might benefit from cancer surveillance. Further studies are essential to accurately assess cancer risk and determine the causative role of *MCM8*/*MCM9* deficiency in cancer predisposition.

## Data Availability

The datasets analyzed during this study are available from the corresponding author on reasonable request.

## Abbreviations

ACMG: American College of Medical Genetics and Genomics
AF: allele frequency
AMP: Association for Molecular Pathology
CADD: Combined Annotation-Dependent Depletion
CI: confidence interval
CRC: colorectal cancer
ERN-GENTURIS: European Reference Network initiative for rare genetic tumor risk syndromes
FFPE: formalin-fixed, paraffin-embedded
ICD10: International Classification of Diseases 10th Revision
ICD-O: International Classification of Diseases for Oncology
MCM8: minichromosome maintenance 8 homologous recombination repair factor
MCM9: minichromosome maintenance 9 homologous recombination repair factor
MMR: mismatch repair
OR: odds ratio
pLoF: predicted loss of function
POI: primary ovarian insufficiency
SBS: single base substitution(s)
SIFT: Sorting Intolerant from Tolerant
SNV: single nucleotide variant
VEP: Ensembl Variant Effect Predictor
VUS: variant of uncertain significance
WES: whole exome sequencing
WGS: whole genome sequencing

## Acknowledgements

The authors sincerely thank all patients and their families for their participation in this study. This research has been conducted using data from UK Biobank (project code 86977), a major biomedical database (www.ukbiobank.ac.uk). Moreover, this research was made possible through access to data in the National Genomic Research Library (project code 1142), which is managed by Genomics England Limited (a wholly owned company of the Department of Health and Social Care). The National Genomic Research Library holds data provided by patients and collected by the NHS as part of their care and data collected as part of their participation in research. The National Genomic Research Library is funded by the National Institute for Health Research and NHS England. The Wellcome Trust, Cancer Research UK and the Medical Research Council have also funded research infrastructure. Finally, this publication and the underlying study have been made possible partly based on data that Hartwig Medical Foundation and the Center of Personalised Cancer Treatment (CPCT) have made available to the study through the Hartwig Medical Database (reference number HMF-DR-288).

---

**Supplementary Figure 1. Phenotype of monoallelic *MCM8/MCM9* variant carriers.** The phenotype is presented for all (**A**) monoallelic *MCM8* and (**B**) monoallelic *MCM9* variant carriers from our case series. Each column represents an individual, while each row corresponds to one of the four primary observed phenotypes: CRC, other type(s) of cancer, hypogonadism, and polyposis. Person IDs are provided below each column. *CRC, colorectal cancer; VUS, variant of uncertain significance*.

**Supplementary Figure 2. Disease onset in monoallelic** MCM8/MCM9 **variant carriers.** The onset of the four primary observed phenotypes (CRC, other type(s) of cancer, hypogonadism, and polyposis) is displayed for each monoallelic *MCM8/MCM9* variant carrier with available age details in our case series. Those without age details were excluded from the analysis. Individuals are ordered by ACMG/AMP classification (pathogenic or likely pathogenic, VUS)^53, 54^ and current age or age at the time of death/lost to follow-up. *ACMG, American College of Medical Genetics and Genomics; AMP, Association for Molecular Pathology; CRC, colorectal cancer; VUS, variant of uncertain significance*.

**Supplementary Figure 3. Monoallelic *MCM8/MCM9* variants mapped onto the respective protein domains.** (**A**) A*MCM8* and (**B**) *MCM9* variants from all monoallelic variant carriers in our case series are mapped onto the domains of the MCM8 and MCM9 proteins, respectively. The fill and color of the diamond symbols correspond to the phenotype of the individual (CRC, other type(s) of cancer, hypogonadism, polyposis) and the ACMG/AMP classification of the variant (pathogenic or likely pathogenic, VUS)^53, 54^, respectively. *ACMG, American College of Medical Genetics and Genomics; AMP, Association for Molecular Pathology; CRC, colorectal cancer; RF, arginine finger; VUS, variant of uncertain significance; WA, Walker A; WB, Walker B; WH, winged-helix; ZF, zinc-finger*.

**Supplementary Figure 4. Mutational signature analysis of metastasized CRCs from monoallelic *MCM8/MCM9* variant carriers in the HMF cancer-specific cohort.** Mutational signature analysis was conducted on metastasized CRCs from monoallelic *MCM8/MCM9* variant carriers in the HMF cancer-specific cohort. The heatmap displays unsupervised hierarchical clustering of the SBS mutational signature profiles. This analysis included tumors from four monoallelic *MCM8* and three monoallelic *MCM9* variant carriers without a second hit in the *MCM8/MCM9* genes. All germline *MCM8/MCM9* variants were classified as VUS per the ACMG/AMP classification for variant interpretation.^53, 54^ The rows represent SBS mutational signatures, while the columns represent individual samples. The identification of SBS mutational signatures was achieved by fitting the counts of SNVs per 96 tri-nucleotide context to the COSMIC signatures^71^, employing the MutationalPatterns tool.^72^ *ACMG, American College of Medical Genetics and Genomics; AMP, Association for Molecular Pathology; HMF, Hartwig Medical Foundation; SBS, single base substitution; SNV, single nucleotide variant; VUS, variant of uncertain significance*.

**Supplementary Figure 5. Mutational signature analysis on TCGA Pan-Cancer Atlas samples harboring somatic** MCM8/MCM9 **mutation(s).** (**A**) Heatmaps showing unsupervised hierarchic clustering of the SBS mutational signature profiles of TCGA Pan-Cancer Atlas tumors with somatic *MCM8/MCM9* mutation(s). Rows represent SBS mutational signatures, while columns represent individual samples. In clusters represented by SBS7a/b (UV damage), SBS2 and SBS13 (APOBEC activity), SBS6, SBS14, SBS15, SBS20, and SBS21 (MMR deficiency), or SBS10a/b (POLE deficiency), the somatic *MCM8/MCM9* variants were likely secondary to other mutational processes.^66, 67, 71, 78, 79^ Tumors with likely pathogenic *MCM8* (n=1) or *MCM9* (n=4) variants in the unexplained clusters were marked by red squares. (**B**) In tumors with likely pathogenic *MCM8* (n=1) or *MCM9* (n=4) variants from the unexplained clusters, shared SBS mutational signatures included SBS22 (aristolochic acid exposure), SBS42 (haloalkane exposure), and SBS54 (sequencing artefact). Notably, SBS mutational signatures associated with MMR deficiency (SBS6, SBS14, SBS15, SBS20, SBS21) were also present in three of these five tumors. However, these could be explained by a likely pathogenic *MLH1* variant and two variants of unknown significance in the *MSH6* gene in one (TCGA-DI-A1BU-01) of these tumors. The TCGA Pan-Cancer Atlas was assessed via cBioPortal for Cancer Genomics (https://www.cbioportal.org/) in February-April, 2023. *ACMG, American College of Medical Genetics and Genomics; AMP, Association for Molecular Pathology; LP, likely pathogenic; mt, mutation; SBS, single base substitution; TCGA, The Cancer Genome Atlas; VUS, variant of uncertain significance; wt, wildtype*.

