## Supplementary Table 1 for "Clinical syndromes linked to biallelic germline variants in *MCM8* and *MCM9*"

| **Supplementary Table 1. ICD10 and ICD-O codes and corresponding phenotypes used to identify cohorts for variant enrichment analysis** | | | | | |
| --- | --- | --- | --- | --- | --- |
| **ICD10 codes** | **Phenotype** | **100K Genomes Project** | | **200K UK Biobank** | |
|  |  | **n**^a^ | **Median age (range)**^b^ | **n**^a^ | **Median age (range)**^b^ |
| K635 | Colonic polyps | 3051 | 70 (5-100) | 9262 | 73 (45-84) |
| K621 | Rectal polyps | 1404 | 69 (5-98) | 5164 | 72 (51-84) |
| D120,D121,D123,D124,D125,D126,D127,D128,D129 | Colorectal adenomas | 2877 | 72 (10-99) | 10297 | 73 (45-84) |
| C180,C181,C182,C183,C184,C185,C186,C187,C188,C189,C19,C20 | Colorectal cancer | 3473 | 73 (17-102 | 3239 | 74 (42-84) |
| C509 | Breast cancer | 4438 | 66 (22-102) | 6783 |  |
| C160, C161, C162, C163, C164, C165, C166, C168, C169 | Gastric cancer | 281 | 70 (18-97) | 481 |  |
| C430, C431, C432, C433, C434, C435, C436, C437, C438, C439 | Melanoma | 714 | 70 (13-102) | 1626 |  |
| C541, C542, C543, C548, C549, C55 | Endometrial cancer | 1107 | 71 (21-99) | 938 |  |
| C56, C561, C562,C563, C569 | Ovarian cancer | 844 |  | 696 |  |
| C530, C531, C538, C539 | Cervical cancer | 135 |  | 145 |  |
| N979 | Female infertility | 434 |  | 495 | 57 (46-72) |
| E283 | Primary Ovarian Insufficiency | 71 |  | 24 |  |
| N46 | Male infertility | 20 |  | 33 |  |
| G400 | Localization-related (focal) (partial) idiopathic epilepsy and epileptic syndromes with seizures of localized onset | 218 | 27 (0-89) | 25 |  |
| G401 | Localization-related (focal) (partial) symptomatic epilepsy and epileptic syndromes with simple partial seizures | 1413 |  | 124 |  |
| G402 | Localization-related (focal) (partial) symptomatic epilepsy and epileptic syndromes with complex partial seizures | 996 |  | 193 |  |
| G403 | Generalized idiopathic epilepsy and epileptic syndromes | 2251 |  | 313 |  |
| G404 | Other generalized epilepsy and epileptic syndromes | 724 |  | 8 |  |
| G405 | Special epileptic syndromes | 81 |  | 31 |  |
| E343 | Short Stature | 994 |  | 9 |  |
| E300 | Delayed Puberty | 129 |  | 0 |  |
| E039 | Hypothyroidism | 3323 | 63 (0-102) | 10997 | 73 (44-84) |
| Q510 | Absent/Infantile Uteri | 13 |  | 0 |  |
| **ICD-O** | **Phenotype** | **100K Genomes Project** | | **200K UK Biobank** | |
|  |  | **n**^a^ | **Median age (range)**^b^ | **n**^a^ | **Median age (range)**^b^ |
| 9061/3  9070/3  9071/3  9100/3  9080/0  9080/1  9085/3  9084/3  9086/3 | Seminoma  Embryonal carcinoma  Yolk sac tumor  Choriocarcinoma  Mature teratoma  Immature teratoma of the yhymus  Mixed germ cell tumor  Teratoma with somatic type malignancies  Germ cell tumor with associated hematological malignancy | 152 |  | 216 |  |
| ^a^ n = number of participants with each phenotype. Some participants had multiple ICD10 codes that were included in our search.  ^b^ The ages of the cases are provided for analyses where variant enrichment could be performed (i.e., at least one homozygous or compound heterozygous case and control was available).  *ICD10, International Classification of Diseases 10th Revision; ICD-O, International Classification of Diseases for Oncology* | | | | | |
