## Supplementary Table 2 for "Clinical syndromes linked to biallelic germline variants in *MCM8* and *MCM9*"

| **Supplementary Table 2. Overview of *MCM8/MCM9* variant carriers meeting pathogenicity-based filtering criteria, including sources** | | | | | | |
| --- | --- | --- | --- | --- | --- | --- |
| **Current study ID** | **Current family ID (only for new cases from outpatient clinic or carriers from literature with updated data)** | **Source** | **Digital Object Identifier (DOI) of source (if applicable)** | **Study ID used in source (if applicable)** | **Germline *MCM8* variant(s)** | **Germline *MCM9* variant(s)** |
| **Biallelic *MCM8* (VUS)** | | | | | | |
| 01143258 |  | Carriers from literature - data from papers only | 10.1016/j.fertnstert.2017.07.015 | V-11 | c.482A>C, p.(His161Pro) |  |
| 02328912 | *MCM8*_04 | Carriers from literature - updated data | 10.1172/jci.insight.140698 | SXS48 | c.692T>A, p.(Ile231Lys); c.994A>G, p.(Thr332Ala) |  |
| 07868858 |  | Carriers from literature - data from papers only | 10.1016/j.ajhg.2022.01.011 | P0281 | c.482A>C, p.(His161Pro) |  |
| 15517012 | *MCM8*_04 | Outpatient clinic |  |  | c.692T>A, p.(Ile231Lys); c.994A>G, p.(Thr332Ala) |  |
| 28590785 |  | Carriers from literature - data from papers only | 10.1016/j.fertnstert.2017.07.015 | V-5 | c.482A>C, p.(His161Pro) |  |
| 39848302 |  | Carriers from literature - data from papers only | 10.1016/j.fertnstert.2017.07.015 | V-9 | c.482A>C, p.(His161Pro) |  |
| 48582770 |  | Carriers from literature - data from papers only | 10.1172/JCI78473 | IV-9 | c.446C>G, p.(Pro149Arg) |  |
| 55456091 |  | Carriers from literature - data from papers only | 10.1172/JCI78473 | IV-6 | c.446C>G, p.(Pro149Arg) |  |
| 71809414 |  | Carriers from literature - data from papers only | 10.1016/j.fertnstert.2017.07.015 | V-10 | c.482A>C, p.(His161Pro) |  |
| 92426276 |  | Carriers from literature - data from papers only | 10.1172/JCI78473 | IV-1 | c.446C>G, p.(Pro149Arg) |  |
| 96184605 |  | Carriers from literature - data from papers only | 10.1016/j.ajhg.2022.01.011 | P0370 | c.482A>C, p.(His161Pro) |  |
| **Biallelic *MCM8* (pathogenic or likely pathogenic)** | | | | | | |
| 01130852 |  | Carriers from literature - data from papers only | 10.1210/jc.2019-00248 | BAB7675 | c.925C>T, p.(Arg309*) |  |
| 08232281 |  | Carriers from literature - data from papers only | 10.1136/jmedgenet-2014-102921 | IV-6 | c.1470_1471insTA, p.(Leu491fs) |  |
| 13147827 |  | Carriers from literature - data from papers only | 10.1002/mgg3.1165 | IV-1 | c.351_354delAAAG, p.(Lys118fs) |  |
| 23867510 |  | Carriers from literature - data from papers only | 10.1136/jmedgenet-2014-102921 | V-1 | c.1954-1G>A, splice acceptor |  |
| 25350218 |  | Carriers from literature - data from papers only | 10.1002/mgg3.1165 | IV-3 | c.351_354delAAAG, p.(Lys118fs) |  |
| 26298807 | *MCM8*_01 | Outpatient clinic |  |  | c.2027A>T, p.(Tyr676Phe); c.1953+1G>C, splice donor |  |
| 38595546 |  | Carriers from literature - data from papers only | 10.1136/jmedgenet-2014-102921 | V-2 | c.1954-1G>A, splice acceptor |  |
| 43675980 |  | Carriers from literature - data from papers only | 10.1136/jmedgenet-2014-102921 | IV-3 | c.1470_1471insTA, p.(Leu491fs) |  |
| 45685855 |  | Carriers from literature - data from papers only | 10.1210/clinem/dgaa155 | IV-2 | c.925C>T, p.(Arg309*) |  |
| 46951971 |  | Carriers from literature - data from papers only | 10.1210/jc.2019-00248 | BAP7675 | c.925C>T, p.(Arg309*) |  |
| 54907964 |  | Carriers from literature - data from papers only | 10.1136/jmedgenet-2014-102921 | IV-2 | c.1470_1471insTA, p.(Leu491fs) |  |
| 57322856 |  | Carriers from literature - data from papers only | 10.1136/jmedgenet-2014-102921 | IV-7 | c.1470_1471insTA, p.(Leu491fs) |  |
| 74081581 |  | Carriers from literature - data from papers only | 10.1136/jmedgenet-2014-102921 | IV-4 | c.1470_1471insTA, p.(Leu491fs) |  |
| 80565678 |  | Carriers from literature - data from papers only | 10.1038/s41431-021-00977-9 | 1 | c.1905+1G>C, splice donor |  |
| 82585307 | *MCM8*_01 | Outpatient clinic |  |  | c.2027A>T, p.(Tyr676Phe); c.1953+1G>C, splice donor |  |
| **Biallelic *MCM9* (VUS)** | | | | | | |
| 31857966 |  | Carriers from literature - data from papers only | 10.1093/hmg/ddaa101 | NOA-144 |  | c.1301A>C, p.(Gln434Pro) |
| 57170234 |  | Carriers from literature - data from papers only | 10.1007/s10815-021-02083-7 | FS0054 |  | c.1291A>G, p.(Met431Val); c.1157C>T, p.(Thr386Met) |
| 58289086 |  | Carriers from literature - data from papers only | 10.1007/s10815-018-1349-4 | FPOI38 |  | c.1784C>G, p.(Thr595Arg); c.905-1G>T, splice acceptor |
| 78493209 | *MCM9*_09 | Outpatient clinic |  |  |  | c.1642C>T, p.(Arg548Trp); c.152A>T, p.(Asn51Ile) |
| 82082774 | *MCM9*_09 | Carriers from literature - updated data | 10.1172/jci.insight.140698 | 011-69294-1 |  | c.1642C>T, p.(Arg548Trp); c.152A>T, p.(Asn51Ile) |
| 88176711 | *MCM9*_08 | Carriers from literature - updated data | 10.1172/jci.insight.140698 | MSS13-1961 |  | c.3425A>G, p.(Lys1142Arg); c.1640T>C, p.(Leu547Pro) |
| **Biallelic *MCM9* (pathogenic or likely pathogenic)** | | | | | | |
| 08014933 | *MCM9*_03 | Carriers from literature - updated data | 10.1016/j.cancergen.2015.10.001 | III-4 |  | c.672_673delGGinsC, p.(Glu225fs) |
| 17276581 | *MCM9*_04 | Carriers from literature - updated data | 10.1038/s41525-021-00242-4 | IV-2 |  | c.1483G>T, p.(Glu495*) |
| 17419906 |  | Carriers from literature - data from papers only | 10.1515/jpem-2020-0590 | P33 |  | c.1732+2T>C, splice donor |
| 30893196 | *MCM9*_05 | Carriers from literature - updated data | 10.3390/jcm12030990 | IV-2 |  | c.394C>T, p.(Arg132*) |
| 37139385 |  | Carriers from literature - data from papers only | 10.1016/j.ajhg.2014.11.002 | AII-6 |  | c.1732+2T>C, splice donor |
| 40636194 |  | Carriers from literature - data from papers only | 10.1210/jc.2019-00248 | BAB10068 |  | c.220C>T, p.(Arg74*) |
| 42545284 |  | Carriers from literature - data from papers only | 10.1515/jpem-2020-0590 | P32 |  | c.1732+2T>C, splice donor |
| 44283624 | *MCM9*_04 | Carriers from literature - updated data | 10.1038/s41525-021-00242-4 | IV-1 |  | c.1483G>T, p.(Glu495*) |
| 54365488 | *MCM9*_06 | Outpatient clinic |  |  |  | c.1720C>T, p.(Arg574*); c.1529-3C>A, splice donor |
| 54601491 |  | Carriers from literature - data from papers only | 10.1111/cge.13803 | POI-02 |  | c.1473dupT, p.(Thr492Tyrfs*4) |
| 54840711 | *MCM9*_05 | Carriers from literature - updated data | 10.3390/jcm12030990 | IV-3 |  | c.394C>T, p.(Arg132*) |
| 67217467 |  | Carriers from literature - data from papers only | 10.1007/s10815-018-1349-4 | FPOI24 |  | c.1651C>T, p.(Gln551*) |
| 69061260 |  | Carriers from literature - data from papers only | 10.1111/cge.13803 | POI-03 |  | c.1473dupT, p.(Thr492Tyrfs*4) |
| 74185426 | *MCM9*_05 | Carriers from literature - updated data | 10.3390/jcm12030990 | IV-5 |  | c.394C>T, p.(Arg132*) |
| 75503761 |  | Carriers from literature - data from papers only | 10.1210/jc.2019-00248 | BAB9435 |  | c.394C>T, p.(Arg132*) |
| 79778808 |  | Carriers from literature - data from papers only | 10.1210/jc.2016-2565 |  |  | c.1651C>T, p.(Gln551*) |
| 80899809 | *MCM9*_05 | Carriers from literature - updated data | 10.3390/jcm12030990 | IV-1 |  | c.394C>T, p.(Arg132*) |
| 80909502 |  | Carriers from literature - data from papers only | 10.1016/j.ajhg.2014.11.002 | AII-4 |  | c.1732+2T>C, splice donor |
| 82941667 |  | Carriers from literature - data from papers only | 10.1016/j.ajhg.2014.11.002 | BII-1 |  | c.394C>T, p.(Arg132*) |
| 91725927 | *MCM9*_10 | Outpatient clinic |  |  |  | c.394C>T, p.(Arg132*) |
| 98796144 | *MCM9*_03 | Carriers from literature - updated data | 10.1016/j.cancergen.2015.10.001 | III-3 |  | c.672_673delGGinsC, p.(Glu225fs) |
| 17290073 | *MCM9*_07 | Outpatient clinic |  |  |  | c.820C>T, p.(Gln274*); c.2237_2238dup, p.(Phe747Ilefs*78) |
| **Monoallelic *MCM8* (VUS)** | | | | | | |
| 19246873 | *MCM8*_04 | Outpatient clinic |  |  | c.692T>A, p.(Ile231Lys) |  |
| 20049146 |  | Carriers from literature - data from papers only | 10.1172/JCI78473 | IV-8 | c.446C>G, p.(Pro149Arg) |  |
| 21357409 |  | Carriers from literature - data from papers only | 10.1186/s12920-020-00813-x | P32 | c.839C>G, p.(Ser280Cys) |  |
| 22206931 |  | Carriers from literature - data from papers only | 10.1210/jc.2019-00248 | BAP7100 | c.89A>C, p.(Lys30Thr); c.1330A>G, p.(Ile444Val) |  |
| 22444712 |  | Carriers from literature - data from papers only | 10.1210/jc.2019-00248 |  | c.89A>C, p.(Lys30Thr) |  |
| 24757813 |  | Carriers from literature - data from papers only | 10.1016/j.fertnstert.2017.07.015 | IV-3 | c.482A>C, p.(His161Pro) |  |
| 27628395 |  | Carriers from literature - data from papers only | 10.1172/JCI78473 | IV-2 | c.446C>G, p.(Pro149Arg) |  |
| 28219291 |  | Carriers from literature - data from papers only | 10.1002/mgg3.1396 | Proband's sister | c.724T>C, p.(Cys242Arg); c.1334C>A, p.(Ala445Asp) |  |
| 28527992 | *MCM8*_03 | Outpatient clinic |  |  | c.482A>G, p.(His161Arg) |  |
| 28664918 |  | Carriers from literature - data from papers only | 10.1172/JCI78473 | IV-5 | c.446C>G, p.(Pro149Arg) |  |
| 33556074 |  | Carriers from literature - data from papers only | 10.1186/s12920-020-00813-x | P28 | c.1565C>T, p.(Thr522Met) |  |
| 33783390 | *MCM8*_04 | Outpatient clinic |  |  | c.692T>A, p.(Ile231Lys) |  |
| 34898570 |  | Carriers from literature - data from papers only | 10.1210/jc.2016-2565 |  | c.1577A>G, p.(Gln526Arg) |  |
| 35155778 | *MCM8*_01 | Outpatient clinic |  |  | c.2027A>T, p.(Tyr676Phe) |  |
| 35922003 |  | Carriers from literature - data from papers only | 10.1016/j.fertnstert.2017.07.015 | V-8 | c.482A>C, p.(His161Pro) |  |
| 37725555 | *MCM8*_04 | Outpatient clinic |  |  | c.692T>A, p.(Ile231Lys) |  |
| 39121178 | *MCM8*_04 | Outpatient clinic |  |  | c.692T>A, p.(Ile231Lys) |  |
| 45566358 |  | Carriers from literature - data from papers only | 10.1172/JCI78473 | III-1 | c.446C>G, p.(Pro149Arg) |  |
| 51426649 |  | Carriers from literature - data from papers only | 10.1210/jc.2019-00248 |  | c.89A>C, p.(Lys30Thr); c.1330A>G, p.(Ile444Val) |  |
| 55019508 |  | Carriers from literature - data from papers only | 10.1016/j.fertnstert.2016.08.018 | 192 | c.950A>T, p.(His317Leu); c.1801_1803del, p.(His601Arg) |  |
| 56285499 |  | Carriers from literature - data from papers only | 10.1016/j.fertnstert.2017.07.015 | V-6 | c.482A>C, p.(His161Pro) |  |
| 57000767 |  | Carriers from literature - data from papers only | 10.1210/jc.2016-2565 |  | c.1561G>A, p.(Asp521Asn) |  |
| 64086299 |  | Carriers from literature - data from papers only | 10.1002/mgg3.1396 | Proband | c.724T>C, p.(Cys242Arg); c.1334C>A, p.(Ala445Asp) |  |
| 65644624 |  | Carriers from literature - data from papers only | 10.1210/jc.2016-2565 |  | c.482A>G, p.(His161Arg) |  |
| 70776442 |  | Carriers from literature - data from papers only | 10.1186/s12920-020-00813-x | P14 | c.1565C>T, p.(Thr522Met) |  |
| 71560890 | *MCM8*_02 | Carriers from literature - updated data | 10.1172/jci.insight.140698 | MSS23-1939 | c.2209G>A, p.(Ala737Thr) |  |
| 76635144 |  | Carriers from literature - data from papers only | 10.1016/j.fertnstert.2017.07.015 | IV-4 | c.482A>C, p.(His161Pro) |  |
| 80115707 |  | Carriers from literature - data from papers only | 10.1210/jc.2016-2565 |  | c.1334G>A, p.(Arg445Gln) |  |
| 91398837 |  | Carriers from literature - data from papers only | 10.1172/JCI78473 | III-2 | c.446C>G, p.(Pro149Arg) |  |
| 93819210 | *MCM8*_04 | Outpatient clinic |  |  | c.692T>A, p.(Ile231Lys) |  |
| 99800629 | *MCM8*_01 | Outpatient clinic |  |  | c.2027A>T, p.(Tyr676Phe) |  |
| **Monoallelic *MCM8* (pathogenic or likely pathogenic)** | | | | | | |
| 19352727 | *MCM8*_01 | Outpatient clinic |  |  | c.1953+1G>C, splice donor |  |
| 23670309 |  | Carriers from literature - data from papers only | 10.1002/mgg3.1165 | III-1 | c.351_354delAAAG, p.(Lys118fs) |  |
| 23830393 |  | Carriers from literature - data from papers only | 10.1136/jmedgenet-2014-102921 | III-1 | c.1470_1471insTA, p.(Leu491fs) |  |
| 35383705 |  | Carriers from literature - data from papers only | 10.1136/jmedgenet-2014-102921 | IV-1 | c.1954-1G>A, splice acceptor |  |
| 39108256 |  | Carriers from literature - data from papers only | 10.1210/jc.2019-00248 |  | c.925C>T, p.(Arg309*) |  |
| 48121907 |  | Carriers from literature - data from papers only | 10.1136/jmedgenet-2014-102921 | IV-5 | c.1470_1471insTA, p.(Leu491fs) |  |
| 48419419 |  | Carriers from literature - data from papers only | 10.1136/jmedgenet-2014-102921 | V-3 | c.1954-1G>A, splice acceptor |  |
| 50513168 |  | Carriers from literature - data from papers only | 10.1210/jc.2019-00248 |  | c.925C>T, p.(Arg309*) |  |
| 53121845 | *MCM8*_05 | Carriers from literature - updated data | 10.1172/jci.insight.140698 | LLS17 | c.351_354delAAAG, p.(Lys118Glufs*5); c.414A>G,  p.(Ile138Met) |  |
| 56752266 |  | Carriers from literature - data from papers only | 10.1002/mgg3.1165 | III-2 | c.351_354delAAAG, p.(Lys118fs) |  |
| 70137960 |  | Carriers from literature - data from papers only | 10.1136/jmedgenet-2014-102921 | III-3 | c.1470_1471insTA, p.(Leu491fs) |  |
| 70350057 |  | Carriers from literature - data from papers only | 10.3390/cancers13040929 | AA3530 | c.876-1delG, ) |  |
| 72640236 |  | Carriers from literature - data from papers only | 10.1210/clinem/dgaa155 | III-2 | c.925C>T, p.(Arg309*) |  |
| 85760408 |  | Carriers from literature - data from papers only | 10.1210/clinem/dgaa155 | III-1 | c.925C>T, p.(Arg309*) |  |
| 86022960 | *MCM8*_01 | Outpatient clinic |  |  | c.1953+1G>C, splice donor |  |
| 95528561 |  | Carriers from literature - data from papers only | 10.1136/jmedgenet-2014-102921 | III-2 | c.1470_1471insTA, p.(Leu491fs) |  |
| 95717575 |  | Carriers from literature - data from papers only | 10.1136/jmedgenet-2014-102921 | IV-2 | c.1954-1G>A, splice acceptor |  |
| 97627350 |  | Carriers from literature - data from papers only | 10.1210/clinem/dgaa155 | IV-1 | c.925C>T, p.(Arg309*) |  |
| **Monoallelic *MCM9* (VUS)** | | | | | | |
| 06583901 |  | Carriers from literature - data from papers only | 10.1016/j.neurobiolaging.2021.12.004 | III-4 |  | c.739C>T, p.(Arg247Trp) |
| 10277227 | *MCM8*_*MCM9*_01 | Carriers from literature - updated data | 10.1172/jci.insight.140698 | MSS2-1941 | c.832C>T, p.(Arg278Cys) | c.3425A>G, p.(Lys1142Arg) |
| 17364805 |  | Carriers from literature - data from papers only | 10.1210/jc.2016-2565 |  |  | c.686T>G, p.(Val229Gly) |
| 17724734 |  | Carriers from literature - data from papers only | 10.1016/j.fertnstert.2019.11.015 | POI-1 |  | c.1423C>T, p.(Leu475Phe) |
| 26113606 | *MCM9*_02 | Carriers from literature - updated data | 10.1172/jci.insight.140698 | NA96-14 |  | c.1915C>G, p.(Leu639Val) |
| 36162360 |  | Carriers from literature - data from papers only | 10.1371/journal.pone.0240795 | POI-25 |  | c.1163C>A, p.(Thr388Asn) |
| 37235179 |  | Carriers from literature - data from papers only | 10.1210/jc.2016-2565 |  |  | c.1784C>G, p.(Thr595Arg) |
| 38753115 |  | Carriers from literature - data from papers only | 10.1016/j.neurobiolaging.2021.12.004 | IV-9 |  | c.739C>T, p.(Arg247Trp) |
| 48901681 |  | Carriers from literature - data from papers only | 10.1016/j.neurobiolaging.2021.12.004 | III-8 |  | c.739C>T, p.(Arg247Trp) |
| 53809122 |  | Carriers from literature - data from papers only | 10.1002/humu.24057 | F39 |  | c.427C>T, p.(Arg143Trp) |
| 54336601 | *MCM9*_06 | Outpatient clinic |  |  |  | c.1529-3C>A, splice donor |
| 58397907 |  | Carriers from literature - data from papers only | 10.1016/j.neurobiolaging.2021.12.004 | III-3 |  | c.739C>T, p.(Arg247Trp) |
| 63437136 |  | Carriers from literature - data from papers only | 10.1016/j.neurobiolaging.2021.12.004 | III-1 |  | c.739C>T, p.(Arg247Trp) |
| 83280371 |  | Carriers from literature - data from papers only | 10.1186/s12920-020-00813-x | P26 |  | c.1330G>C, p.(Val444Leu) |
| 90757541 |  | Carriers from literature - data from papers only | 10.1210/jc.2016-2565 |  |  | c.905-1G>T, splice acceptor |
| **Monoallelic *MCM9* (pathogenic or likely pathogenic)** | | | | | | |
| 00367545 | *MCM9*_03 | Carriers from literature - updated data | 10.1016/j.cancergen.2015.10.001 | III-2 |  | c.672_673delGGinsC, p.(Glu225fs) |
| 00369720 |  | Carriers from literature - data from papers only | 10.1515/jpem-2020-0590 |  |  | c.1732+2T>C, splice donor |
| 00882217 |  | Carriers from literature - data from papers only | 10.1016/j.ajhg.2014.11.002 | AI-1 |  | c.1732+2T>C, splice donor |
| 02099761 |  | Carriers from literature - data from papers only | 10.1016/j.ajhg.2014.11.002 | AI-2 |  | c.1732+2T>C, splice donor |
| 07793607 | *MCM9*_03 | Carriers from literature - updated data | 10.1016/j.cancergen.2015.10.001 | II-1 |  | c.672_673delGGinsC, p.(Glu225fs) |
| 14948286 |  | Carriers from literature - data from papers only | 10.1210/jc.2016-2565 |  |  | c.2011G>T, p.(Glu671*) |
| 15964832 |  | Carriers from literature - data from papers only | 10.1210/jc.2019-00248 |  |  | c.220C>T, p.(Arg74*) |
| 24996014 |  | Carriers from literature - data from papers only | 10.1016/j.ajhg.2014.11.002 | BII-2 |  | c.394C>T, p.(Arg132*) |
| 25080587 | *MCM9*_03 | Carriers from literature - updated data | 10.1016/j.cancergen.2015.10.001 | III-1 |  | c.672_673delGGinsC, p.(Glu225fs) |
| 26472597 |  | Carriers from literature - data from papers only | 10.1016/j.ajhg.2014.11.002 | BI-1 |  | c.394C>T, p.(Arg132*) |
| 29508735 | *MCM9*_01 | Carriers from literature - updated data | 10.1172/jci.insight.140698 | NA41-1 |  | c.1987dupT, p.(Ser663Phefs*36) |
| 35173643 |  | Carriers from literature - data from papers only | 10.1016/j.ajhg.2014.11.002 | AII-3 |  | c.1732+2T>C, splice donor |
| 40559303 | *MCM9*_04 | Carriers from literature - updated data | 10.1038/s41525-021-00242-4 | IV-3 |  | c.1483G>T, p.(Glu495*) |
| 41229278 | *MCM9*_03 | Carriers from literature - updated data | 10.1016/j.cancergen.2015.10.001 | II-2 |  | c.672_673delGGinsC, p.(Glu225fs) |
| 42152259 |  | Carriers from literature - data from papers only | 10.1210/jc.2019-00248 |  |  | c.220C>T, p.(Arg74*) |
| 42750756 |  | Carriers from literature - data from papers only | 10.1210/jc.2019-00248 |  |  | c.394C>T, p.(Arg132*) |
| 49294952 | *MCM9*_04 | Carriers from literature - updated data | 10.1038/s41525-021-00242-4 | III-2 |  | c.1483G>T, p.(Glu495*) |
| 58731687 |  | Carriers from literature - data from papers only | 10.1016/j.ajhg.2014.11.002 | AII-2 |  | c.1732+2T>C, splice donor |
| 62507727 |  | Carriers from literature - data from papers only | 10.1210/jc.2019-00248 |  |  | c.394C>T, p.(Arg132*) |
| 71823158 | *MCM9*_03 | Carriers from literature - updated data | 10.1016/j.cancergen.2015.10.001 | I-1 |  | c.672_673delGGinsC, p.(Glu225fs) |
| 76457125 |  | Carriers from literature - data from papers only | 10.1016/j.ajhg.2014.11.002 | BII-4 |  | c.394C>T, p.(Arg132*) |
| 77887094 |  | Carriers from literature - data from papers only | 10.1016/j.ajhg.2014.11.002 | BI-2 |  | c.394C>T, p.(Arg132*) |
| 82416835 |  | Carriers from literature - data from papers only | 10.1515/jpem-2020-0590 |  |  | c.1732+2T>C, splice donor |
| 92396046 | *MCM9*_04 | Carriers from literature - updated data | 10.1038/s41525-021-00242-4 | IV-4 |  | c.1483G>T, p.(Glu495*) |
| 94130086 |  | Carriers from literature - data from papers only | 10.1210/jc.2016-2565 |  |  | c.2011G>T, p.(Glu671*) |
| 95282910 | *MCM9*_04 | Carriers from literature - updated data | 10.1038/s41525-021-00242-4 | III-3 |  | c.1483G>T, p.(Glu495*) |
| 98962558 | *MCM9*_05 | Carriers from literature - updated data | 10.3390/jcm12030990 | IV-6 |  | c.394C>T, p.(Arg132*) |
| 99284187 | *MCM9*_06 | Outpatient clinic |  |  |  | c.1720C>T, p.(Arg574*) |
| 59356494 | *MCM9*_07 | Outpatient clinic |  |  |  | c.820C>T, p.(Gln274*) |
| 63108002 | *MCM9*_07 | Outpatient clinic |  |  |  | c.2237_2238dup, p.(Phe747Ilefs*78) |
