## Supplementary figures and images for "Clinical syndromes linked to biallelic germline variants in *MCM8* and *MCM9*"

### Supplementary Figure 1

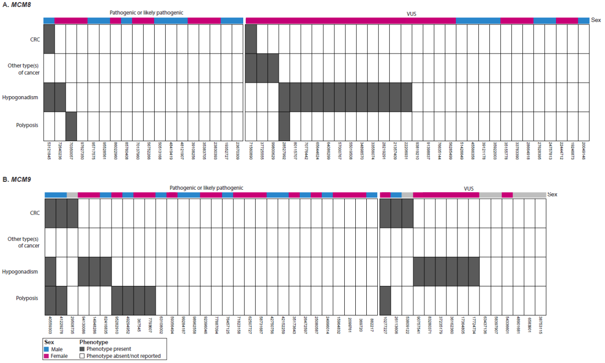

### Supplementary Figure 2

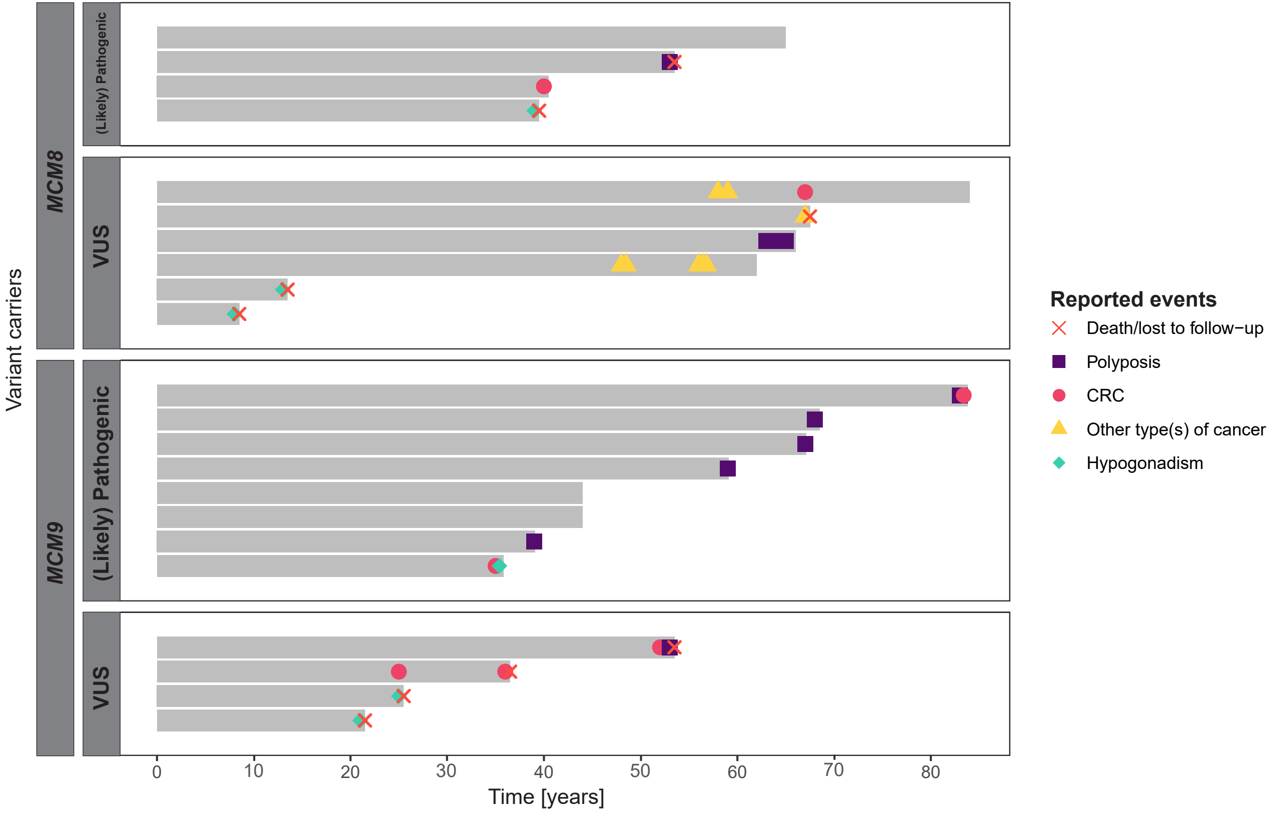

### Supplementary Figure 3

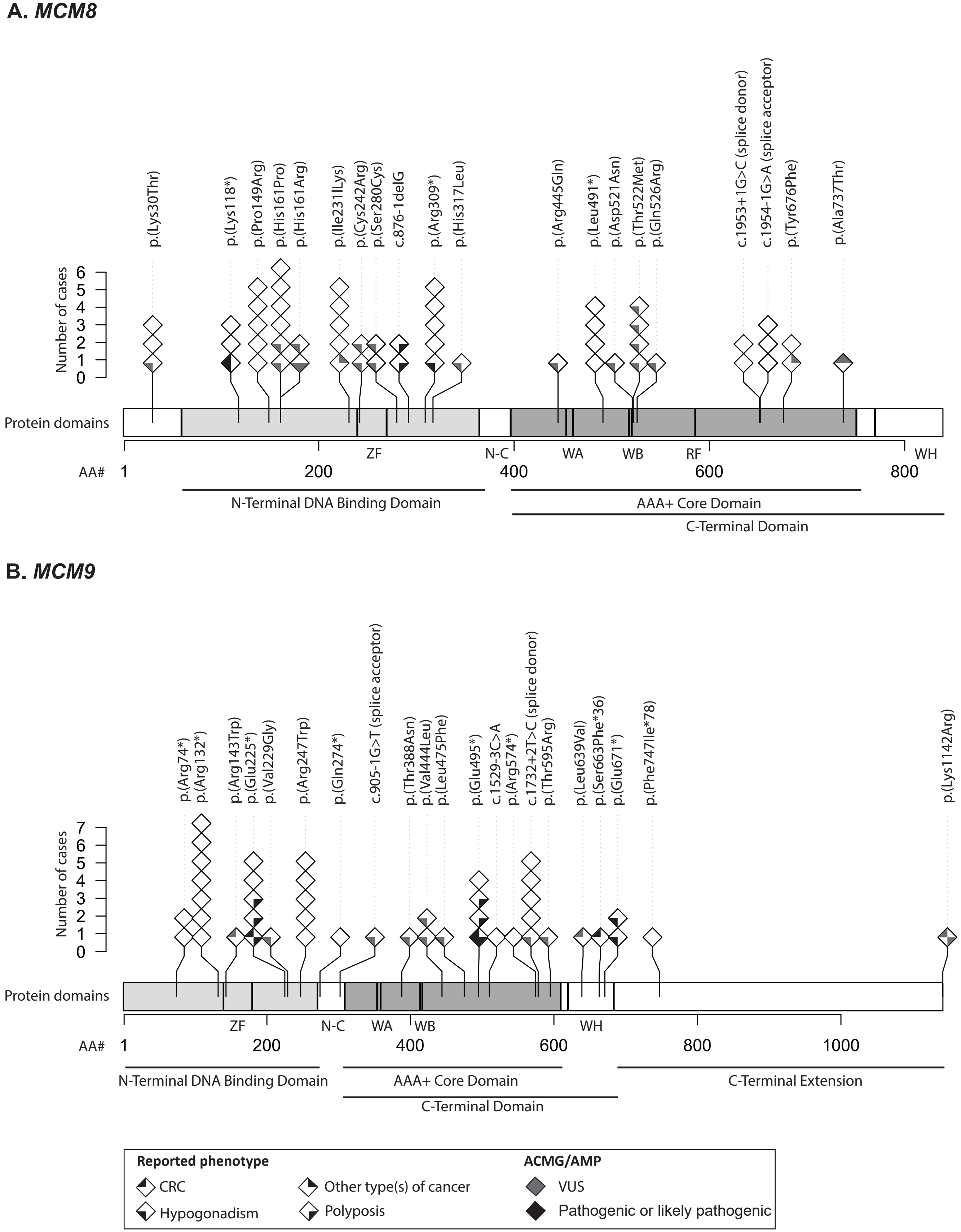

### Supplementary Figure 4

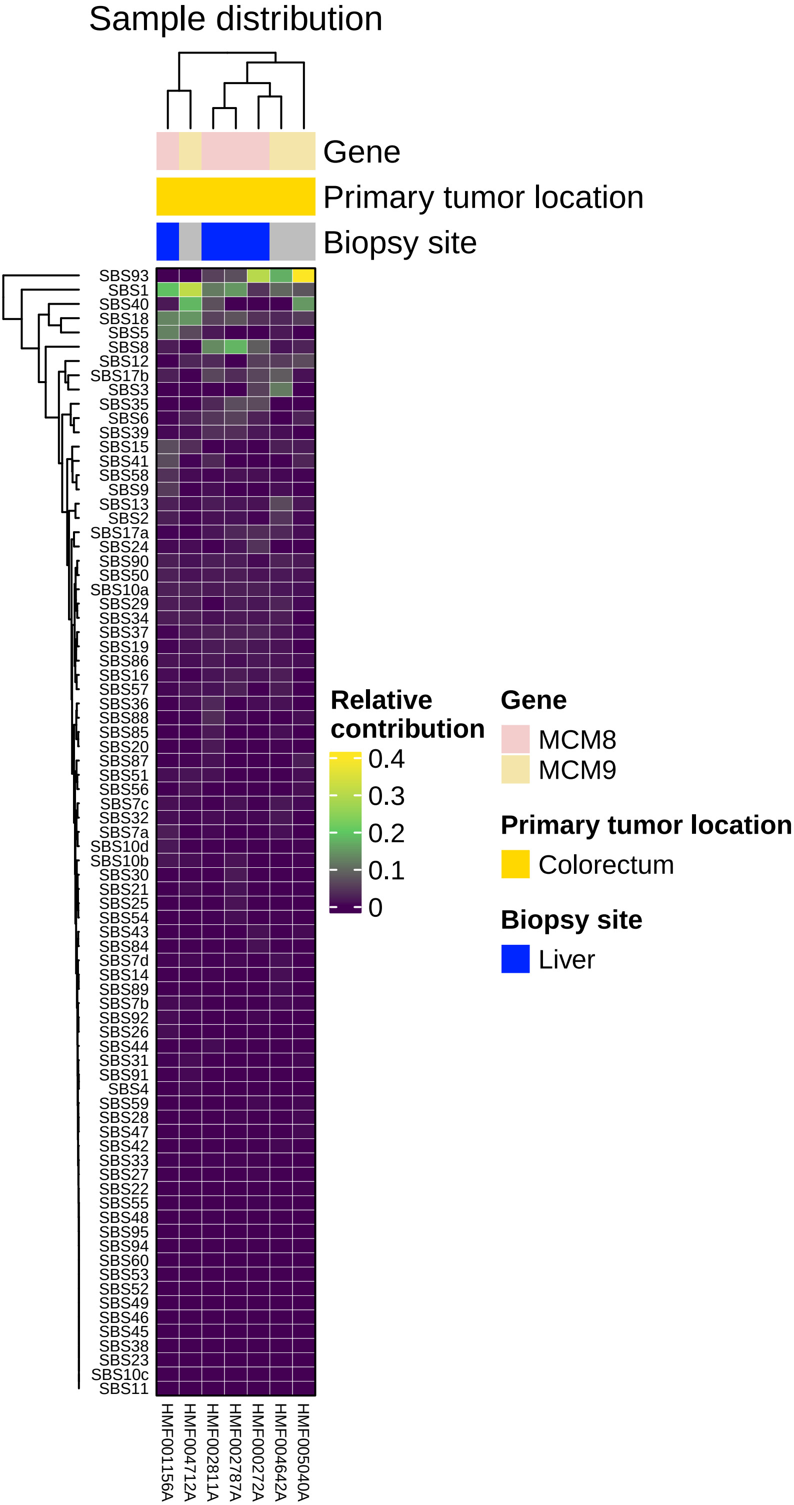

### Supplementary Figure 5

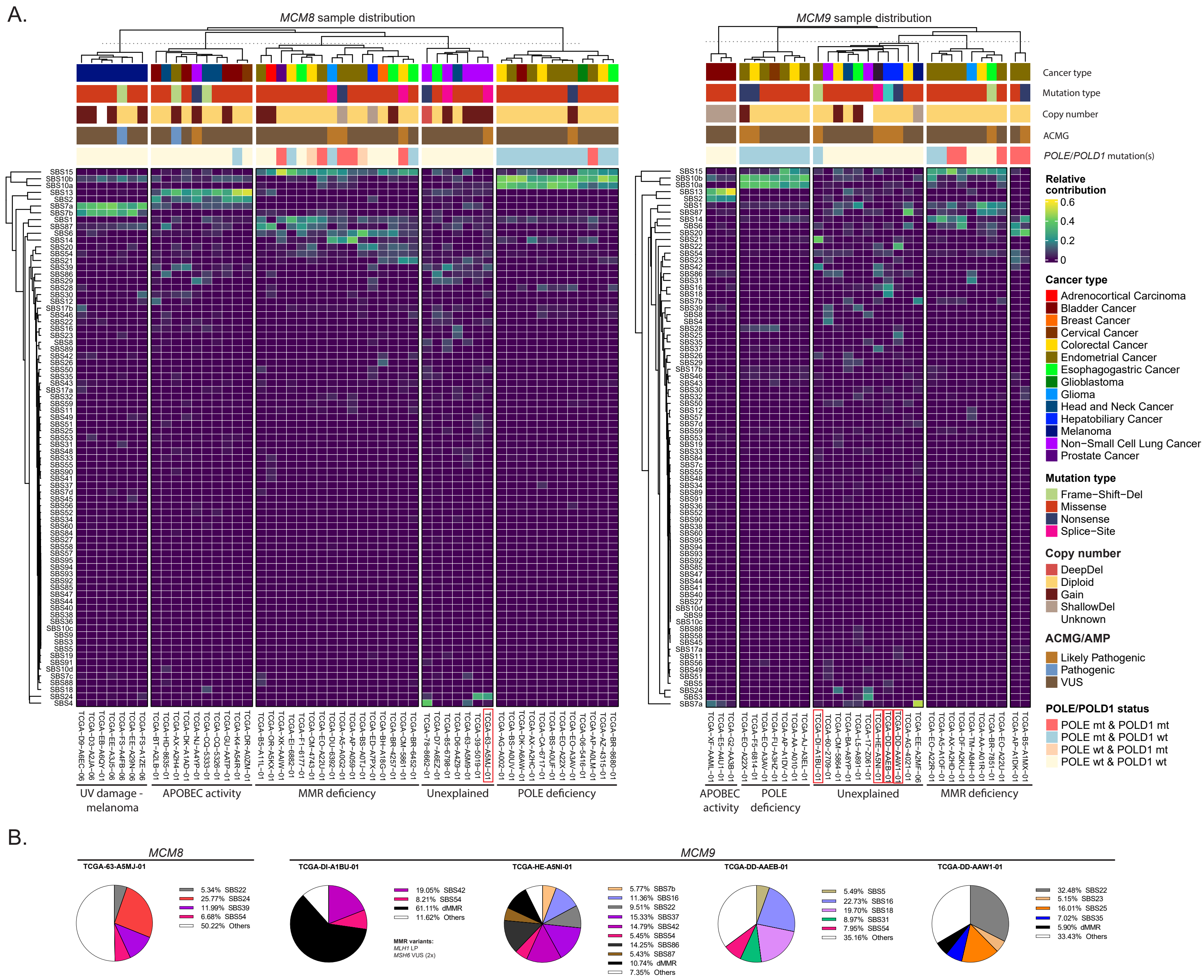
